# Threshold-free genomic cluster detection to track transmission pathways in healthcare settings

**DOI:** 10.1101/2020.09.26.20200097

**Authors:** Shawn E. Hawken, Rachel D. Yelin, Karen Lolans, Robert A. Weinstein, Michael Y. Lin, Mary K. Hayden, Evan S. Snitkin, for the CDC prevention Epicenters program

**Affiliations:** Departments of Microbiology & Immunology, University of Michigan Medical School, Ann Arbor, MI, USA; Internal Medicine Division of Infectious Diseases University of Michigan Medical School, Ann Arbor, MI, USA; Department of Internal Medicine, Division of Infectious Diseases, Rush University Medical Center, Chicago, IL, USA

## Abstract

**Background:** Over the past decade, whole-genome sequencing (WGS) has become the gold standard for tracking the spread of infections in healthcare settings. However, a critical barrier to the routine application of WGS for infection prevention is the lack of reliable criteria for determining if a genomic linkage is consistent with transmission.

**Methods:** Here, we sought to understand the genomic landscape in a high-transmission healthcare setting by performing WGS on 435 carbapenem-resistant *Enterobacterales* (CRE) isolates collected from 256 patients through admission and biweekly surveillance culturing of virtually every hospitalized patient over a 1-year period.

**Findings:** Our analysis revealed that the standard approach of employing a single-nucleotide variant (SNV) threshold to define transmission would lead to both false-positive and false-negative inferences. False positive inferences were driven by the frequent importation of closely related strains, which were presumably linked via transmission at a connected healthcare facility. False negative inferences stemmed from the diversity of colonizing populations being spread among patients, with multiple examples of hypermutator strains emerging within patients and leading to putative transmission links separated by large genetic distances. Motivated by limitations of an SNV threshold, we implemented a novel threshold-free transmission cluster inference approach whereby each of the 234 acquired CRE isolates were linked back to the imported CRE isolate with which it shared the most variants. This approach yielded clusters that varied in levels of genetic diversity but were highly enriched in patients sharing epidemiologic links. Holistic examination of clusters highlighted extensive variation in the magnitude of onward transmission stemming from the more than 100 importation events and revealed patterns in cluster propagation that could inform improvements to infection prevention strategies.

**Interpretation:** Overall, our results show how the integration of culture surveillance data into genomic analyses can overcome limitations of cluster detection based on SNV-thresholds and improve our ability to track pathways of pathogen transmission in healthcare settings.

**Funding:** CDC U54 CK000481, CDC U54 CK00016 04S2. S.E.H was supported by the University of Michigan NIH Training Program in Translational Research T32-GM113900 and the University of Michigan Rackham pre-doctoral fellowship.

**Research in context:** *Evidence before this study:* We searched PubMed for studies published before May 1, 2021, with no start date restriction, with the search “transmission AND whole-genome AND (snp OR snv) AND (cut-off OR threshold) NOT (SARS-CoV-2 OR virus or HIV)”. We identified 18 reports that used whole genome sequencing to study transmission, primarily in healthcare settings. Several of these studies attempted to identify optimal single nucleotide variant (SNV) cutoffs for delineating transmission. These studies were all single-site and had only partial sampling of healthcare facilities. Moreover, even when the same species was considered, different optimal SNV thresholds were reported.

*Added value of this study:* To understand the limitations of an SNV threshold approach for tracking transmission we leveraged a data set that comprised admission and every-other-week CRE surveillance culturing for every patient entering a hospital over the course of one year. By performing genomic analysis of 435 isolates from the 256 CRE colonized patients we systematically demonstrated pitfalls with the use of SNV thresholds for transmission inference that stem from the importation of closely related strains from connected healthcare facilities, variation in genetic heterogeneity of colonizing populations and uneven evolutionary rates of CRE strains colonizing patients. We went on to implement an alternative approach for tracking transmission in healthcare facilities that relies on genetic context, instead of genetic distance to group patients into intra-facility transmission clusters. We applied this approach to our CRE genomes and demonstrated that the resultant transmission clusters are strongly enriched in patients with spatiotemporal overlap, and that clusters can be interrogated to identify putative targets to interrupt transmission.

*Implications of all the available evidence:* Advances in the speed and economy of genome sequencing are making it increasingly feasible to perform routine sequencing to track transmission in healthcare settings. However, a critical barrier to these efforts is the lack of clear criteria for inferring transmission that generalizes to diverse strains of healthcare pathogens and that are robust to variation in organism prevalence and differences in connectivity of local healthcare networks. Here, we show that by combining genome sequencing with surveillance data that healthcare transmission can be inferred in a threshold-free manner. The success of this approach in a setting with high importation and transmission rates bodes well for its generalizability to less challenging healthcare settings.

## Introduction

Healthcare-associated infections (HAIs) are a major threat to patient safety.^1^ Despite increased attention to infection prevention in healthcare settings, cross-transmission between hospitalized patients still occurs, suggesting that there remain poorly understood pathways of nosocomial transmission.^2^ Integration of genomics with traditional hospital epidemiological investigations has proved powerful in the identification of routes of HAI transmission ^3–5^, bringing hope that broad deployment of sequencing can direct improvements in infection prevention practices. However, a major barrier to the use of genomics to track the spread of infections is a lack of clear, robust and generalizable criteria to assess whether two patients are linked by transmission.

The current gold standard for inferring transmission is the imposition of a single-nucleotide variant (SNV) threshold, above which transmission in the facility is deemed implausible, and below which transmission is deemed likely^6–8^. However, SNV thresholds have been shown to be imprecise, particularly for tracking the spread of the successful epidemic strains that are responsible for the majority of antibiotic resistance in healthcare settings^9–13^. One source of false positive transmission inferences using SNV thresholds are patients harboring closely related strains due to transmission at a connected healthcare facility. Evidence of this being a significant issue comes from the observation of closely related strains circulating in healthcare facilities that share many patients^9,11,14^. Thus, in a setting of regional endemicity, the accuracy of an SNV threshold is likely to be greatly impacted by transmission rates at connected healthcare facilities.

In addition to false positive inferences, there is abundant evidence that the use of SNV thresholds can lead to false negative inferences, whereby actual transmission events are excluded due to higher-than-expected SNV distances. A well-established source of large SNV distances among true transmission pairs is genetic variation that arises in a patient during prolonged asymptomatic colonization^12,15,16^. In particular, with the common practice of sequencing single isolates from a patient’s colonizing population, it becomes possible for patients linked by transmission to harbor isolates as genetically distant as any two members of the source patient’s colonizing population. Given that many healthcare-associated pathogens are capable of colonizing patients for many months and even years^17,18^, it is likely common for individuals to harbor genetically diverse colonizing populations. Evidence in support of this issue comes from both the observation of large genetic variation in patient’s colonizing populations^16,19^, and the fact that the most accurate SNV thresholds for recent transmission are consistent with diversity that accumulates over years^12,13^.

While the above issues with SNV thresholds are well-known, it remains unclear how significant a barrier they represent to accurate transmission inference in healthcare settings. Moreover, given the need for easily interpretable criteria to guide infection prevention, there currently is a lack of viable alternatives. Here, we sought to understand how prominent the theoretic limitations to SNV thresholds are and develop an alternative strategy that relies on genetic context instead of genetic distance to identify patients linked by transmission. By leveraging a unique dataset comprising admission and bi-weekly surveillance of virtually every patient entering a hospital over the course of a year, we show that the potential issues leading to misattribution of transmission when using SNV thresholds are prominent for carbapenem-resistant *Enterobacterales* (CRE) in an endemic setting. We go on to show by taking advantage of surveillance culture data, that we were able to link patients acquiring CRE back to the imported strains with which they share the most variants, and thereby identify patients linked by transmission in a threshold-free manner.

## Methods

### Study design, clinical setting, and sample collection

The current study was reviewed and approved by both the institutional review boards at Rush University Medical Center and the University of Michigan. Informed consent was waived. Detailed information regarding the study design, intervention bundle and data collection are available in Hayden et. al 2015.^20^ Briefly, a 1-year intervention to prevent CRE colonization and infection took place from 2012-2013 in a Chicago long-term acute care hospital (LTACH). In particular, the intervention focused on *Klebsiella pneumoniae* harboring the blaKPC carbapenemase (KPC-Kp), as this was the dominant CRE type in the region. The intervention included rectal surveillance swab culture-based screening using the direct ertapenem disk method followed by PCR confirmation of *bla*-KPC,^20,21^ of all LTACH patients for KPC-Kp rectal colonization at LTACH admission and every two weeks thereafter until a patient received a positive test (94% adherence), physical separation of KPC-Kp-positive and KPC-Kp-negative patients by placing KPC-Kp-positive patients in ward cohorts (91% adherence), daily chlorhexidine bathing of all patients in the LTACH and a hand hygiene campaign.^20–22^

### Patient surveillance categories

Patients were grouped into categories based on surveillance culture results. Patients who were either positive at the start of the study or within three days of LTACH admission were considered potential sources of KPC-Kp importation and onward transmission within the LTACH. Patients who were KPC-Kp-negative on their first surveillance culture, and then KPC-Kp-positive after day three of admission, were assumed to have acquired KPC-Kp in the facility. If a patient’s first surveillance sample was collected more than three days after admission and was positive for KPC-Kp, the patient was also assumed to have acquired KPC-Kp in the facility for the purposes of the transmission cluster detection algorithm (see below). When an admission-positive patient acquired an additional KPC-Kp strain (as evidenced by multi-locus sequence type (MLST) inferred from WGS data) during their stay this was termed a secondary acquisition, and such isolates from admission positive patients were eligible to be included as acquisition isolates for transmission cluster detection.

### Whole genome sequencing & genome processing

Glycerol stocks containing unique morphologies of KPC-Kp isolates were stored at -80°C prior to cultivation on LB agar for DNA isolation.^20,21^ DNA was extracted with the MoBio PowerMag Microbial DNA kit and prepared for sequencing on an Illumina MiSeq instrument using the NEBNext Ultra kit and sample-specific barcoding. Library preparation and sequencing were performed at the Center for Microbial Systems at the University of Michigan or the University of Michigan Sequencing Core. Quality of reads was assessed with FastQC,^23^ and Trimmomatic^24^ was used for trimming adapter sequences and low-quality bases. In total, 462 isolates were sequenced, with the 435 isolates from 256 unique patients passing QC being used in downstream analyses.

### Data availability

Sequence data are available under BioProject PRJNA603790^25^.

### Identification of single nucleotide variants

SNV calling was performed as in Han *et al*.^26^ The variant calling pipeline can be found at https://github.com/Snitkin-Lab-Umich/variant_calling_pipeline. To summarize, raw reads were mapped to the MLST specific reference genomes listed in **Supplementary Table 1** using bwa. Variant calling was performed with samtools.^27,28^ MLST specific reference genomes were chosen in order to maximize the number of potential shared variants detected among genomes in the study.

### Whole-genome sequence analyses

Whole-genome sequence alignments containing core and non-core variant positions were used to generate pairwise (genome by genome) single-nucleotide variant (SNV) matrices and shared-variant matrices, interrogate mutational biases, query SNVs and indels in mismatch repair genes, and construct phylogenetic trees for transmission cluster detection and descriptions of genomic variants. PanIsa was used to detect insertion sequences in bam files containing WGS alignments.^29^ All whole-genome sequence analyses were performed in R version 3.6.1.

### Transmission cluster detection

Transmission cluster detection using a SNV threshold-free approach was performed on isolates from MLSTs that were present in at least two patients including at least one acquisition patient, as this represents molecularly plausible cross-transmission within the LTACH during the study (**Table 1**). Whole-genome sequence alignments including core and non-core genome variant positions were used to generate maximum parsimony phylogenetic trees, pairwise shared variant matrices and SNV distance matrices for each MLST-specific alignment. Transmission clusters were detected by probing phylogenetic trees for the maximum subtree (**Figure 2A)** containing admission or study-start isolates from a single patient that were collected prior to or at the same time as acquisition isolates in the subtree. We required cluster-defining subtrees to be supported by at least one unique subtree-defining variant that was not found elsewhere in the phylogeny.

**Table 1:** Distribution of KPC-Kp strains isolated from colonized LTACH patients.

| MLST | †Number of isolates | Number of patients | Number of admission positive/study-start isolates | Number of patients with admission positive/study-start isolates | Number of acquisition isolates | Number of acquisition patients | Number of admission positive patients with acquisitions occurring afterwards | Number of acquisition patients potentially acquiring colonization from a plausible source | *Potential cross-transmission link in the LTACH |
| --- | --- | --- | --- | --- | --- | --- | --- | --- | --- |
| 13 | 63 | 37 | 25 | 18 | 38 | 22 | 18 | 15 | yes |
| 14 | 7 | 5 | 2 | 1 | 5 | 4 | 1 | 2 | yes |
| 15 | 17 | 11 | 1 | 1 | 16 | 10 | 1 | 7 | yes |
| 16 | 47 | 32 | 15 | 15 | 32 | 20 | 15 | 17 | yes |
| 20 | 6 | 6 | 1 | 1 | 5 | 5 | 1 | 4 | yes |
| 36 | 1 | 1 | 1 | 1 | 0 | 0 | 0 | 0 | no |
| 134 | 1 | 1 | 1 | 1 | 0 | 0 | 0 | 0 | no |
| 193 | 2 | 2 | 2 | 2 | 0 | 0 | 0 | 0 | no |
| 258 | 271 | 177 | 101 | 83 | 170 | 104 | 83 | 86 | yes |
| 327 | 6 | 4 | 0 | 0 | 6 | 4 | 0 | 0 | yes |
| 834 | 2 | 1 | 2 | 1 | 0 | 0 | 0 | 0 | no |
| 874 | 7 | 5 | 4 | 3 | 3 | 2 | 2 | 1 | yes |
| 883 | 1 | 1 | 1 | 1 | 0 | 0 | 0 | 0 | no |
| 950 | 3 | 1 | 0 | 0 | 3 | 1 | 0 | 0 | no |
| Novel | 1 | 1 | 1 | 1 | 0 | 0 | 0 | 0 | no |

\*Potential cross-transmission link in LTACH during study inferred by at least two patients with isolate of the same MLST and at least one patient acquiring colonization with an isolate of the same MLST.
†Isolate total represents isolates with quality WGS data; 27 samples were excluded from the 462 total isolates obtained due to poor sequence quality.

Multiple admission/study-start positive patients were permitted in clusters if they shared at least one unique variant with the other cluster members. Clusters with no admission/study-start isolates (acquisition isolate only clusters) were permitted if no subtree existed that included an admission/study-start isolate, or if expansion of the subtree would break a valid cluster. Only clusters that contained isolates from at least two patients and at least one acquisition isolate were considered valid transmission clusters for downstream analyses.

### Analysis of location data

Location data were abstracted from patient bed traces, i.e. patient bed and room location(s) over time. Spatiotemporal overlap explanations for cross-transmission between patients in clusters were defined as patients being in the same location (e.g. facility, ward or room) at the same time during the period between when a putative donor patient in the cluster was last negative for the isolate up until and including the day the recipient tested positive for the isolate. The last-negative date was chosen as a conservative bound for the earliest time acquisition could have occurred in order to account for acquisitions occurring between biweekly sampling dates. Sequential exposure was evaluated for the same timeframe, but restricted to patients in the same location separated by time, where the putative donor had been in a location first and the recipient later occupied the same location in the window between their transition from negative to positive surveillance, and no spatiotemporal exposure between donors in the cluster and the recipient could explain the recipients’ acquisition.

### Statistical analysis

Two-sample Kolomogorov-Smirnov tests were used to test for a statistical difference in pairwise SNV distance distribution between admission and acquisition isolates. Multinomial tests were used to determine significant biases in mutational frequencies in transmission cluster isolates compared to overall frequencies of mutation types among all isolates of the same MLST collected in the study. The Wilcoxon rank sum test was used to detect differences in intra-patient and intra-cluster SNV distances between admission and acquisition isolates. Permutation tests were used to evaluate enrichment in spatiotemporal and sequential exposures between patients in transmission clusters.

## Results

### High KPC-Kp colonization burden among hospitalized patients due to high rates of importation and transmission

On the first day of the yearlong study, all patients in the hospital underwent rectal surveillance cultures to detect colonization with *Klebsiella pneumoniae* harboring the bla-KPC carbapenemase (KPC-Kp), which identified 51 colonized patients. During the rest of the study, admission surveillance detected another 77 patients who were positive within three days of first admission, and therefore presumed to have imported KPC-Kp into the facility. In addition, 128 patients were presumed to have acquired KPC-Kp colonization in the hospital either due to having at least one negative surveillance culture prior to a positive or having been in the facility for more than three days before having a surveillance culture taken. While acquisition and importation fluctuated over time **(Figure 1B)** the overall colonization prevalence was consistently high, averaging 32% over the course of the year **(Figure 1A)**. Classification of the 435 isolates from 256 KPC-Kp colonized patients by MLST revealed that 62% of the isolates obtained during the study belonged to ST258, the major epidemic lineage of KPC-Kp in the U.S. (**Table 1**), with six other lineages showing evidence of intra-facility spread as inferred from their detection in at least one importation and acquisition culture (**Table 1, Figure S1**). The seven lineages with putative in-hospital transmission links were imported between one and 83 times each and were the source of between two and 104 acquisitions over the course of the study (**Table 1**). Patients harboring these strains had extensive shared time in the facility (**Figure 2, Figure S2**), demonstrating the complexity of deciphering transmission chains in the facility.

**Figure 1:**
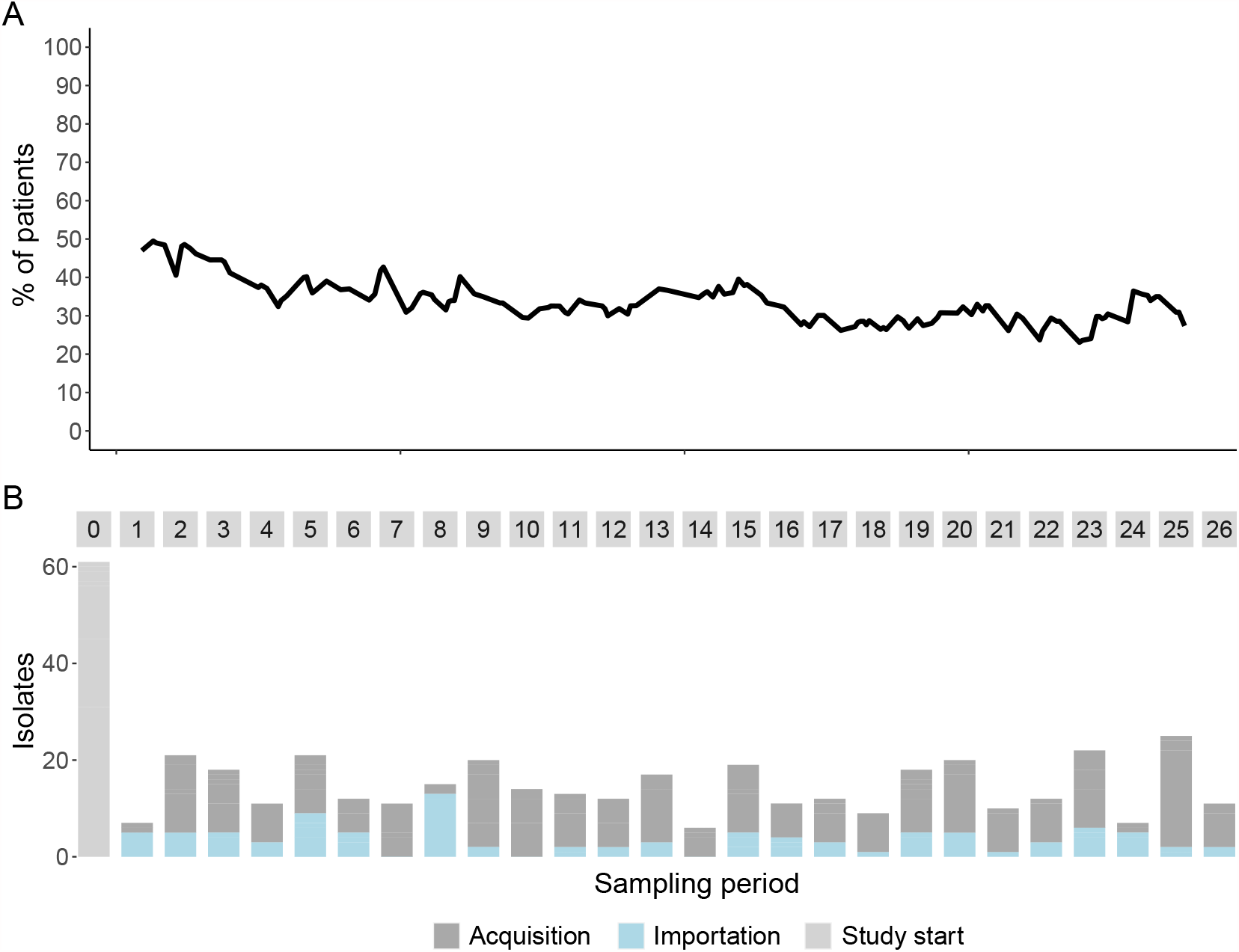
Endemicity of KPC-Kp in the LTACH is due to extensive importation and acquisition. **A**. KPC-Kp prevalence (black line) defined as number of patients presently in the LTACH who are or ever had been surveillance positive for at least one KPC-Kp isolate during the study divided by the number of patients in the facility throughout the 1-year study. **B**. Isolates obtained through bi-weekly rectal surveillance culturing of LTACH patients. Grey boxes indicate the study start (time 0) and every two 14-day surveillance periods (28 days) throughout the study. Bars indicate the KPC-Kp isolates collected at the beginning of the study, for which importation or acquisition status is not known (light grey, study start), within 3 days of the patient first entering the facility (blue, importation), or after negative surveillance or >3 days after ever being in the LTACH during the study (dark grey, acquisition).

**Figure 2:**
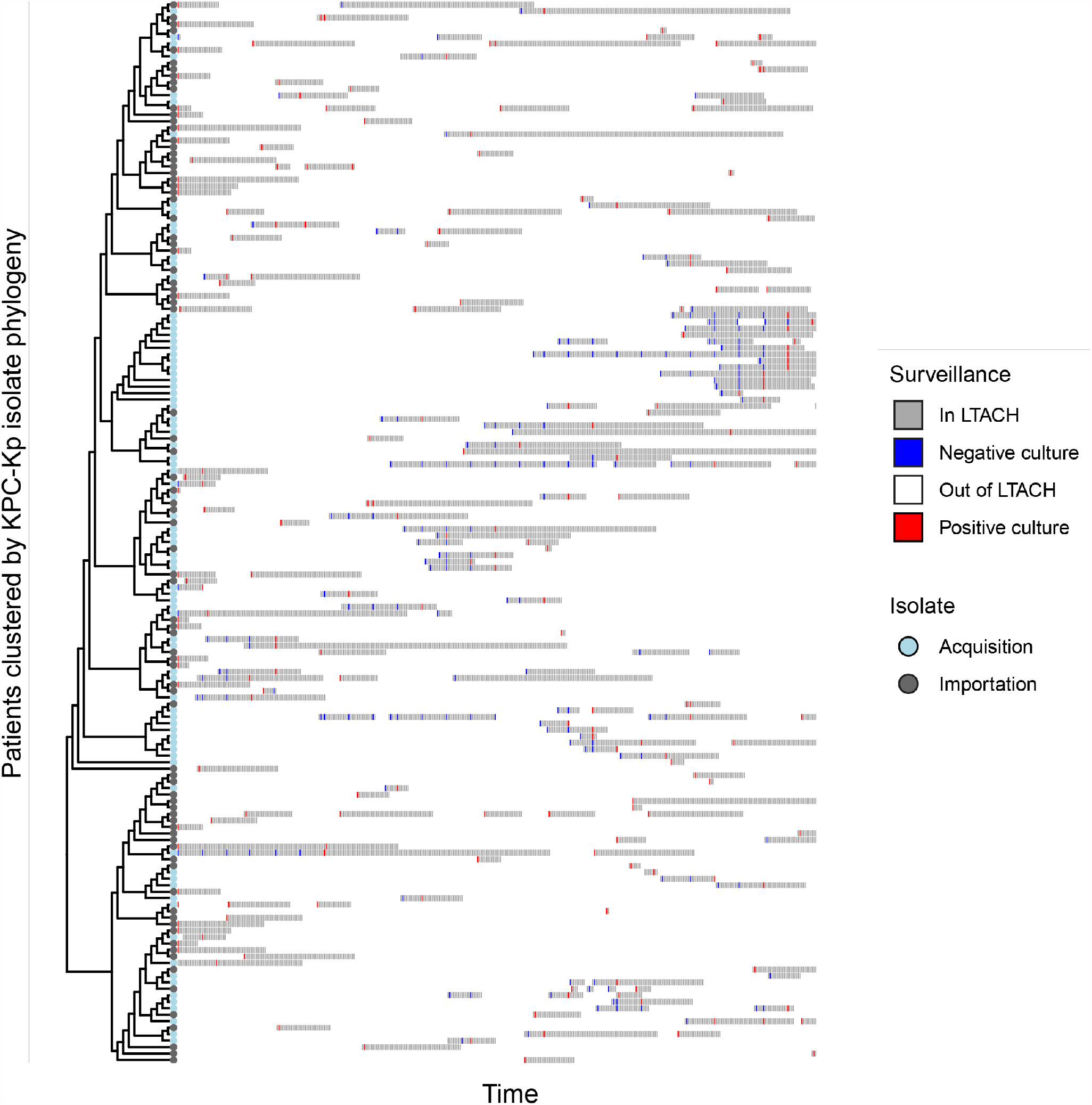
Tracing transmission links within the LTACH is complicated by both extensive importation and acquisition of related strains of KPC-Kp, and patients with shared time exposures in the LTACH. Patient bed trace showing surveillance culture data for patients who tested positive for KPC-Kp strain ST258, at any point during the study. The order of patients on the Y-axis is by the phylogenetic relationship between isolates collected from them. Grey bars indicate patients are in the LTACH, white indicates the patient is out outside of the facility, red indicates positive surveillance culture, blue indicates negative surveillance culture dates. Plots illustrating other MLSTs detected during the study are shown in **Figure S2**.

### Frequent importation of closely related strains prevents imposition of a single SNV threshold to identify patients linked by intra-facility transmission

We next applied the increased resolution of WGS to discern which patients were linked by cross-transmission. First, we examined the potential of applying an SNV threshold to identify patients with isolates linked by cross-transmission that occurred in the hospital during the study. As alluded to above, one type of error associated with the use of SNV thresholds are false positive transmission inferences due to the importation of closely related strains that are linked by prior transmission between patients in a connected healthcare facility. To assess the impact of this in our data, we compared the distribution of genetic distances among pairs of isolates imported into the facility, to genetic distances among pairs including an imported and acquired isolate. **Figure 3** shows that these two distributions are completely overlapping, and that it is commonplace for imported isolates to be related to one another by small genetic distances. Thus, in this high-prevalence endemic setting, there is no SNV threshold that accurately discriminates patients linked by transmission in the facility during the current admission versus during an earlier admission at a connected facility.

**Figure 3:**
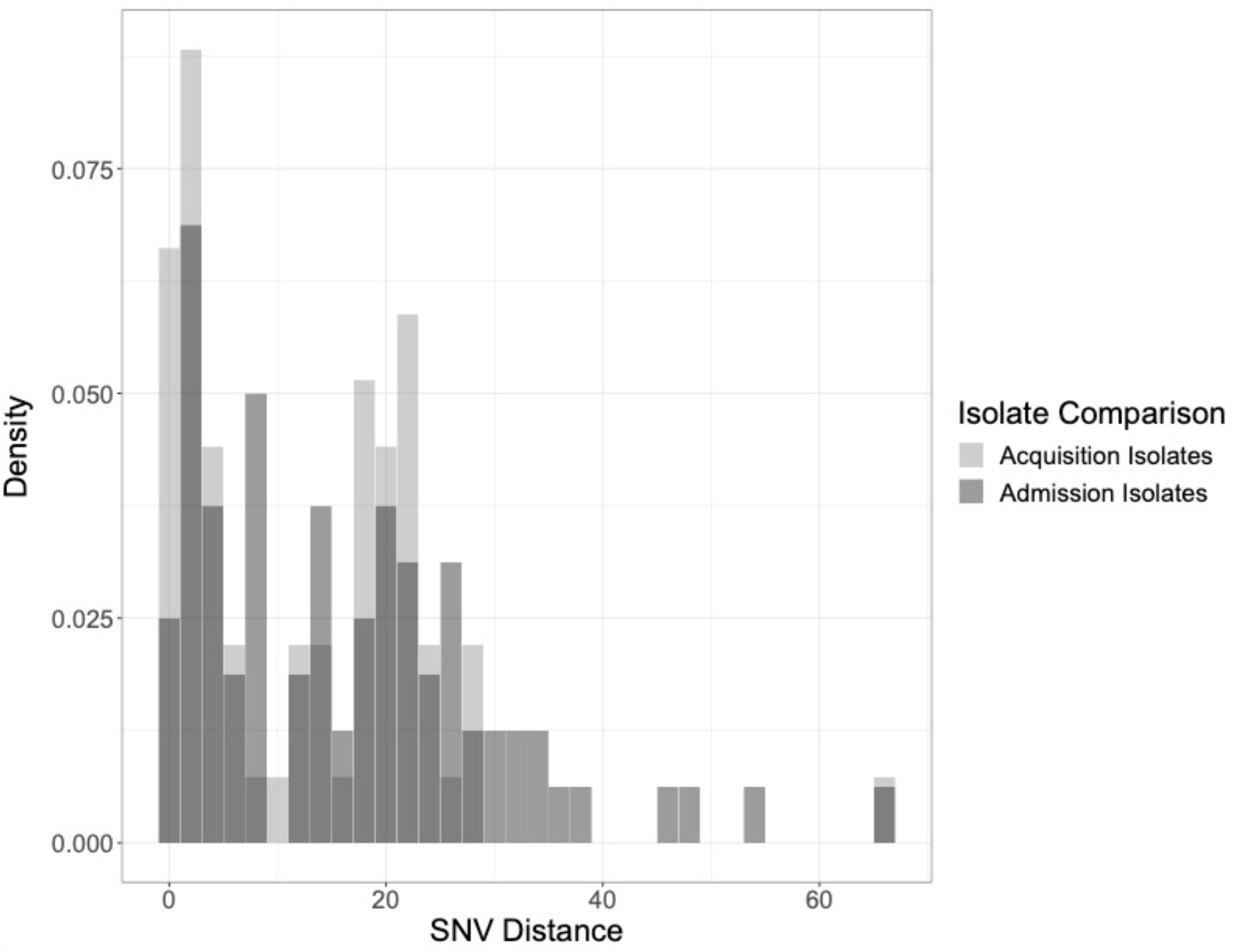
There is no single-nucleotide variant threshold that distinguishes isolates acquired in the LTACH from isolates that are imported by admission-positive patients. Comparison of minimum pairwise SNV distances between the closest related imported isolate and acquired or imported isolates. X-axis indicates SNV distance, Y-axis indicates density (i.e. normalized count) of KPC-Kp isolates from ST258. Light grey bars indicate the minimum distance between isolates collected from patients who acquired KPC-Kp colonization after being in the LTACH >3 days during the admission-positive isolates. Dark grey bars indicate the minimum distance between isolates collected from patients who were positive upon admission to the LTACH. Note that study-start positive patients who were KPC-Kp-positive on the first day of the study, who represent a mixture of recent and prior colonization, were considered admission-positive for this analysis so that acquisitions derived from those transmission chains could be linked. Two-sample Kolmogorov-Smirnov test for differences in the distribution of pairwise SNV distances, p-value = 0.277 (ST258), and p-value=0.39 (non-ST258), 0.81 (all MLSTs combined).

### Grouping KPC-Kp isolates into transmission clusters based on genetic context allows for threshold-free transmission inference

To circumvent the lack of a discriminatory SNV threshold, we sought to apply an approach that relies of genetic context, instead of genetic distance, to identify patients linked by transmission in the facility. To this end we took advantage of our comprehensive knowledge of which patients imported and acquired KPC-Kp and applied an algorithm whereby phylogenetic clusters were identified that grouped each acquisition isolate with the most closely related admission or study-start isolate taken from a patient earlier in the study. In essence, this approach attempts to track each acquisition isolate back to the patient who imported it into the facility by identifying the importation isolate with which it shares a most recent common ancestor (**Figure S4**). Our implementation also allowed for multiple admission positive patients to be included in a cluster if they were equally genetically plausible, and for clusters with no admission positive patient if no suitable option existed (See **Methods**).

Application of this genomic cluster detection method yielded 49 putative transmission clusters grouping a median of 3 (range 2-14) patients into clusters representing at least one acquisition event and at least two patients (**Figure 4**). For 40 of the 49 clusters all cluster members could be traced back to a putative importation isolate (**Table 2**). For 7 of the 9 clusters that do not have an admission positive patient as the first cluster member, there was spatiotemporal overlap explaining all intra-cluster transmission events, indicating that only the source of the cluster was not identified (**Table 2**). In addition to the 49 transmission clusters, there were 18 (14%) patients colonized with KPC-Kp at study start or on admission surveillance screen whose isolates were not linked with a transmission cluster, and therefore presumed to not be associated with onward transmission of KPC-Kp.

**Table 2.**
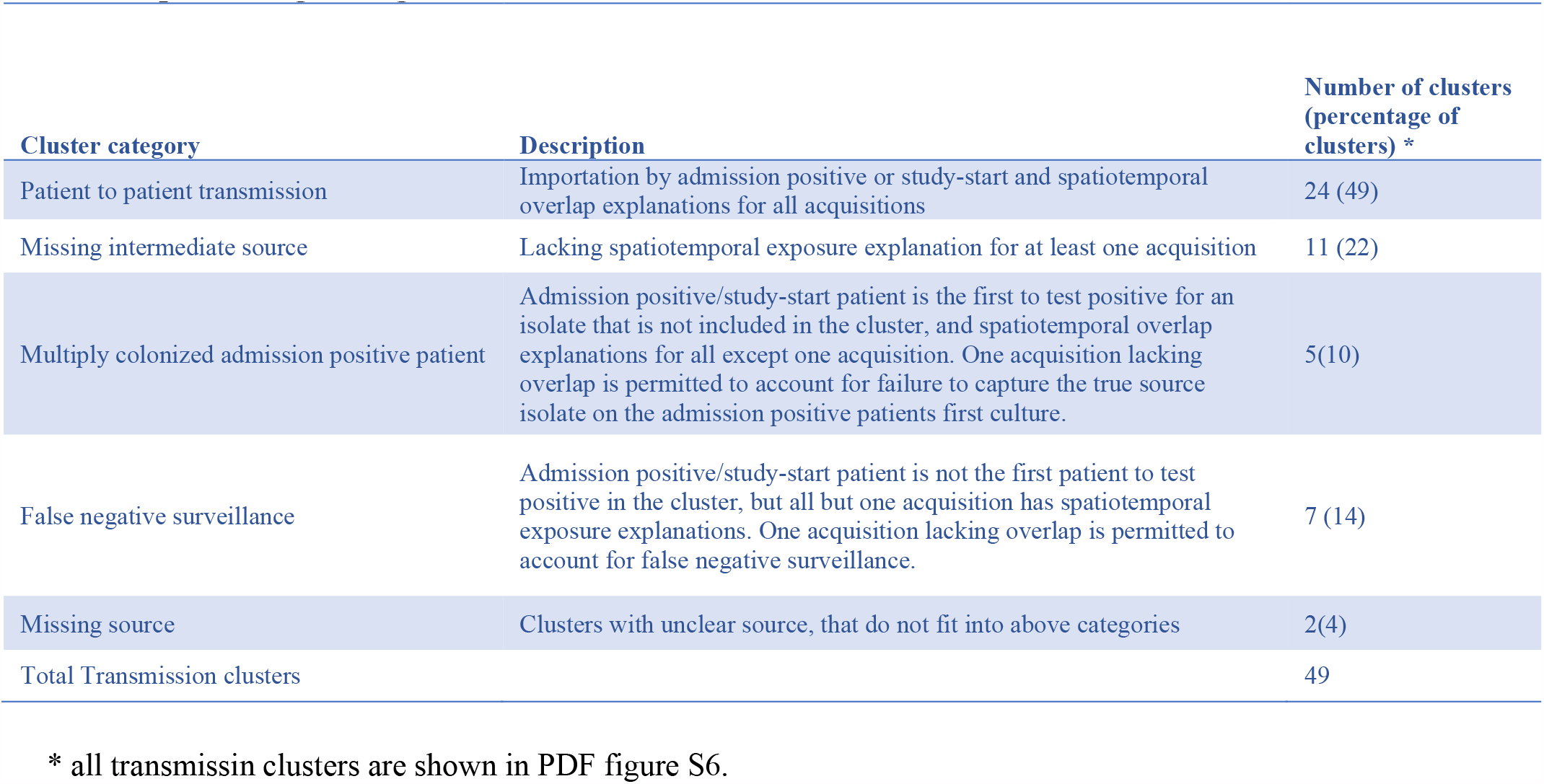
Epidemiologic categorization of transmission clusters.

**Figure 4:**
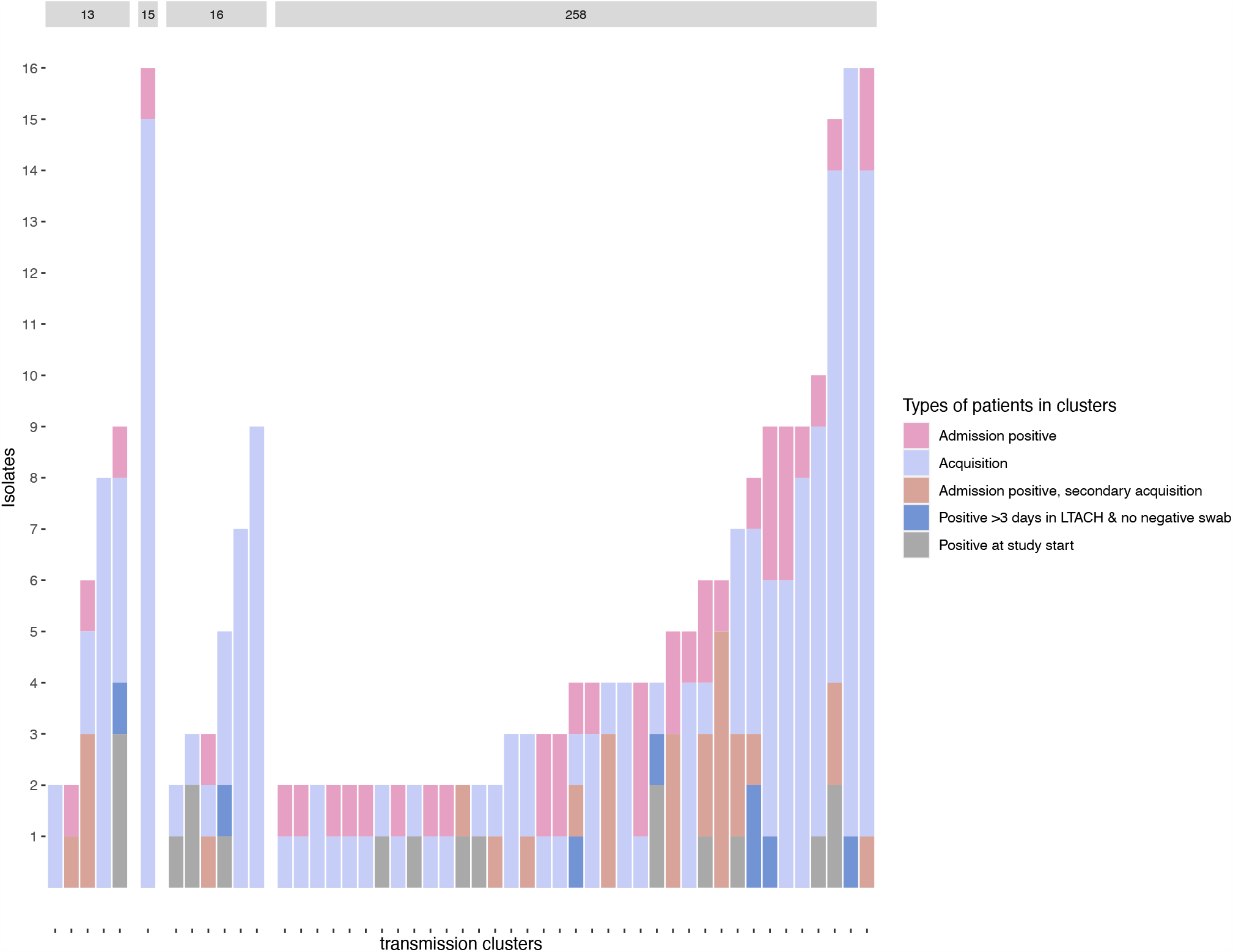
Transmission clusters detected with method based on shared genomic variants and robust surveillance data links the majority of KPC-Kp acquisitions during the study. Distribution of isolates and patients in the 49 transmission clusters detected with genomic method described in supplemental Figure 4. Each column represents isolates from one cluster. Admission positive patients (pink) are patients whose isolate in the cluster was obtained within 3 days of first admission to the facility. Periwinkle indicates isolates obtained from acquisition patients who first acquired KPC-Kp colonization more than 3 days after first admission to the LTACH. Tan indicates isolates from admission positive patients that were collected >3 days after admission to the LTACH, indicating either prolonged colonization or secondary strain acquisition in the LTACH. Blue indicates patients who were first positive after being in the LTACH for >3 days, but from whom no negative swab was collected prior to first KPC-Kp detection. Grey bars indicate patients who were positive on the first day of the study. Bar across top of figure indicates MLST of isolates.

### Transmission clusters detected with an SNV-threshold independent approach harbor a wide range of genetic diversity

Having identified transmission clusters without imposing an SNV threshold, we next sought to characterize the range and variation of genetic diversity associated with putative intra-facility transmission. Calculation of the intra-cluster diversity revealed that the maximum number of SNVs separating pairs of isolates in identified clusters ranged from zero to 153, with a median of nine SNVs. While the majority of clusters varied by small genetic distances, nine clusters (18 %) had larger SNV distances (greater than 30 SNVs) (**Figure 5A**). One source of large SNV distances could be the improper inclusion of admission positive patients who are not the true source of the transmission cluster. Indeed, for two of the nine clusters where the intra-cluster genetic diversity was >30 SNVs (**Figure 5A**, cluster 258_21, 258_108), we found that two admission positive patients were included, one of which was genetically distant from other cluster members. This suggests that while genetic context was unable to discriminate between two putative source patients for these clusters, that genetic distance was informative.

**Figure 5:**
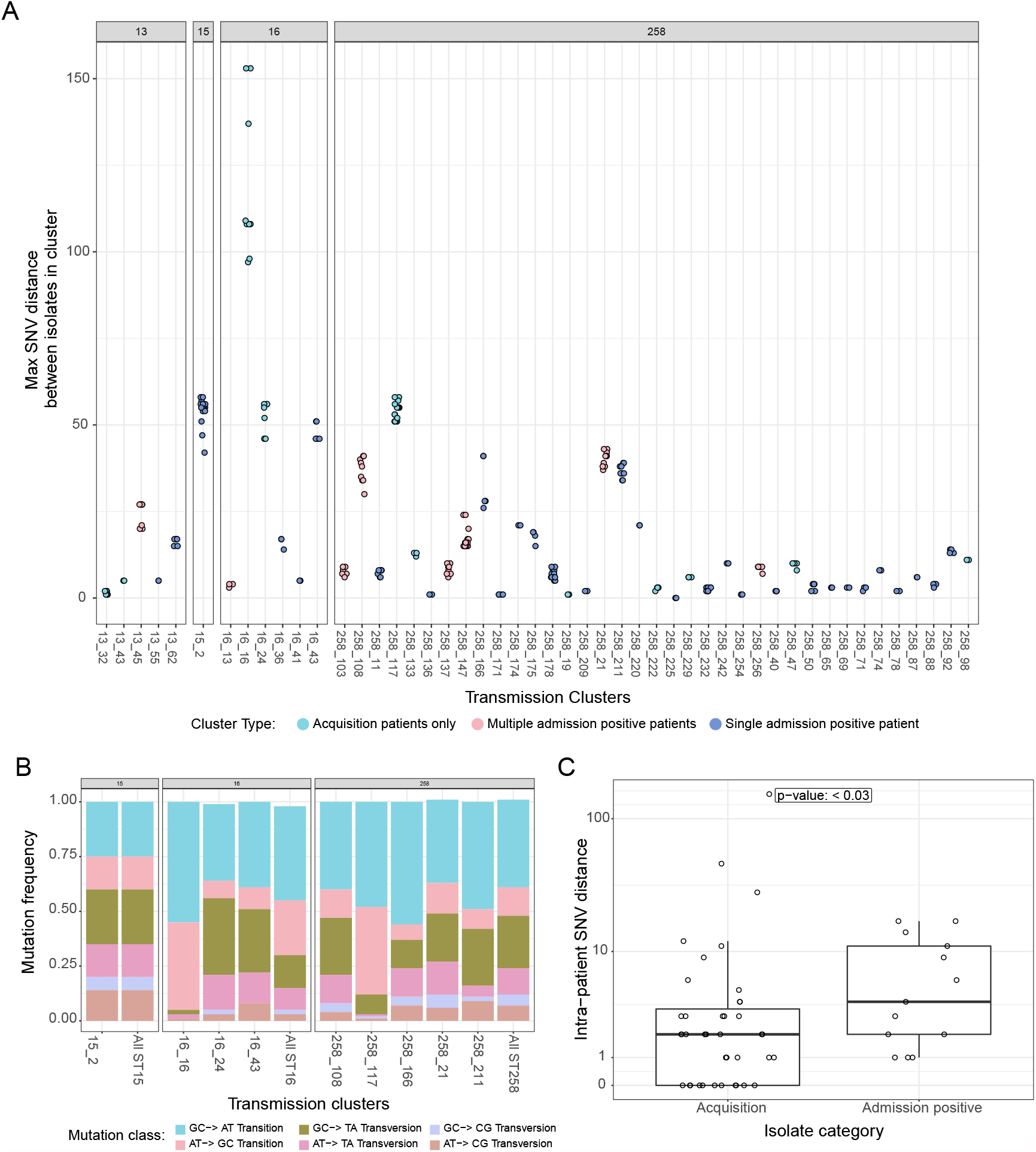
Elevated genetic diversity in transmission clusters is attributable to prolonged colonization and emergence of hypermutator strains. Grey bars indicate the MLST of the isolates in transmission clusters. **A**. Maximum pairwise SNV distance distinguishing isolates from the same cluster. Colors indicate whether the cluster has only acquisition patients, multiple admission positive/study-start patients or a single admission positive patient. **B**. Observed frequencies in mutational classes across isolates included in each transmission cluster among clusters with a maximum pairwise SNV distance of 30 SNV or greater. Bars on the right of each MLST group indicate the overall population frequency of mutational classes among members of that MLST in the study population. Statistically significant skews in mutational frequencies were observed in clusters with large SNV distances (16_16 and 258_117), supporting the role of mismatch repair mutations (e.g. hypermutators). **C**. Maximum intra-patient intra-cluster genetic diversity among admission positive and acquisition patients. Intra-patient intra-cluster genetic diversity is greater among isolates from admission positive patients (Wilcoxon rank sum test, p-value < 0.03).

A second source of elevated inter-patient SNV distances could be the accumulation of genetic variation during prolonged asymptomatic colonization, and potential propagation of this variation via transmission. In support of this, we observed a distribution of intra-patient diversity among both admission positive and acquisition patients who contributed multiple isolates to a cluster (**Figure 5C**). Moreover, we observed a significantly greater intra-patient diversity among admission-positive patients, (Wilcoxon rank sum test p-value < 0.03), supporting the role of prolonged colonization driving intra-patient diversity (**Figure 5C**).

In our examination of intra-patient diversity, we also observed several cases of extreme SNV distances that were inconsistent with previously reported evolutionary rates for KPC-Kp^11,30^. We hypothesized that these large distances could be due to the emergence of hyper-mutator phenotypes, as has been reported for other commensal and pathogenic bacteria.^31^ Genomic signatures of hypermutators include specific mutational biases as well as disruption of DNA mismatch repair genes, which can lead to greater than expected number of mutations in a given time. Analyses of these genomic signatures revealed that the transmission clusters with the largest numbers of intra cluster SNVs (cluster 16_16 -153 SNVs and cluster 258_117 - 58 SNVs), **Figure 5B**) had clear skews in their mutational frequencies (multinomial test, p-value < 0.05) as well as a large insertion in *mutS* in isolates from one of the clusters (cluster 258_117).

### Overlaying location information on transmission clusters reveals pathways of transmission

While the high KPC-Kp colonization prevalence hindered the ability to perform contact tracing in the absence of genomic data, we hypothesized that by examining patterns of overlap within clusters we could gain insight into how KPC-Kp spread among patients in the facility. To this end, we first examined spatiotemporal overlap among cluster members during putative transmission windows (i.e. between negative and positive surveillance cultures), and found that 81%, 66% and 8.5% of KPC-Kp acquisitions could be explained by overlap with another cluster member at the level of facility, floor and room, respectively. Compared to random groups of patients of the same size and patient type distribution (i.e. numbers of admission-positive and acquisition patients), actual transmission clusters were strongly enriched for these spatiotemporal overlaps among patients (**Figure 6** permutation tests, P-value < 0.001, all locations). In contrast to strong evidence for transmission between patients overlapping in space and time, we found little evidence for persistent environmental contamination as a source of transmission, with only 8.5% of acquisitions across clusters explained by sequential exposure to the facility, 4.7% for sequential exposure to a ward, and 0.78% for sequential exposure to a room (**Figure 6**). Of special note given previous reports of sinks as a vehicle for longitudinal transmission, examination of sequential exposures to common rooms among individual cluster patients revealed only a single patient (**Figure S5** cluster 258_175, patient 174), whose sequential exposure to a room previously occupied by another patient from their cluster was the only exposure detected that could explain their acquisition.

**Figure 6:**
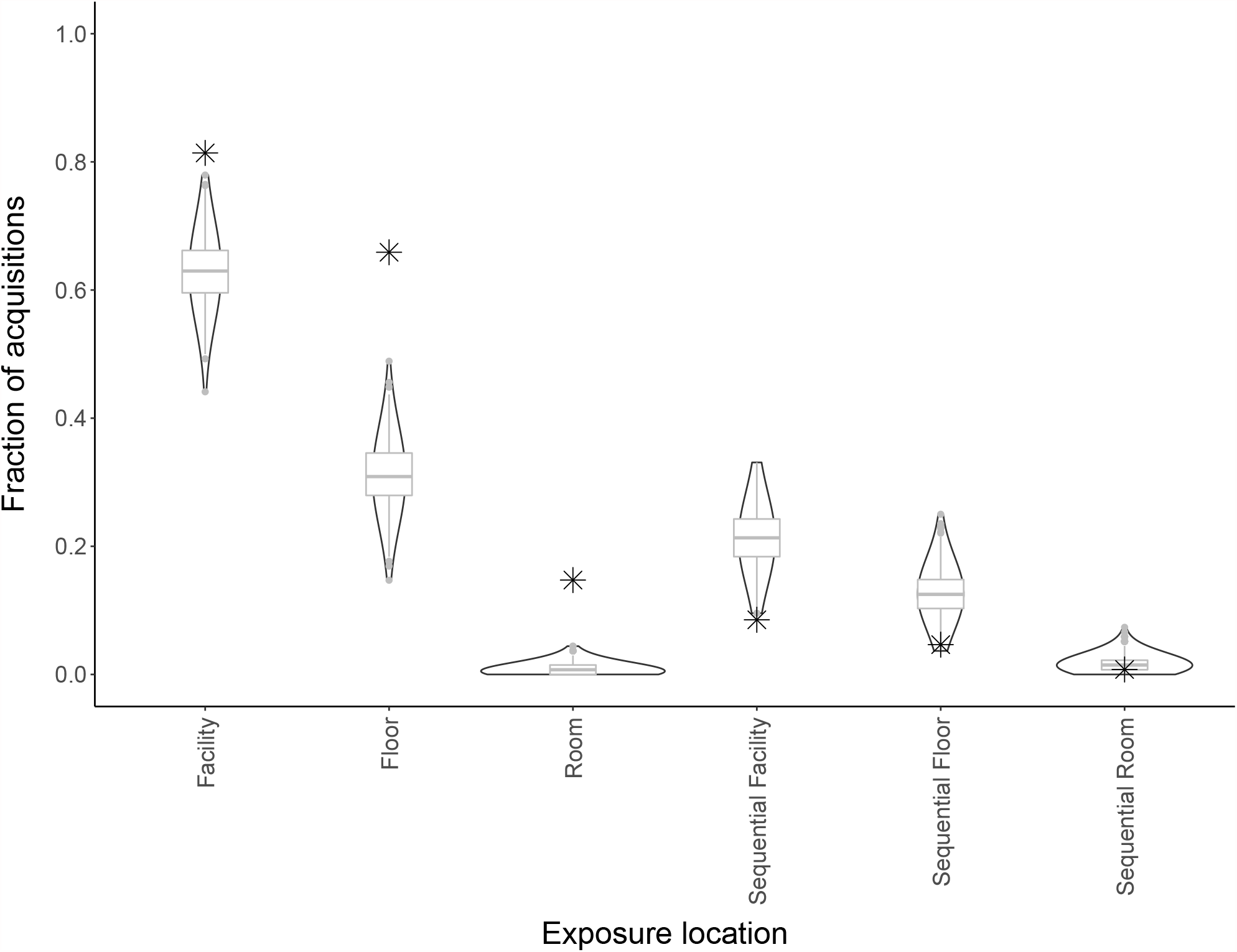
Epidemiologic exposures within transmission clusters point to frequent acquisition of an isolate from outside of a patient’s room location (i.e., ward or facility) and infrequent acquisition linked to sequential occupation of of same room, ward, or facility. X-axis indicates locations, Y axis indicates fraction of acquisitions in transmission clusters that could be attributed to putative donor and recipient (acquisition) patients in the cluster being in the same place at the same time (spatiotemporal exposure) or in the same place separated by time after a donor had left that location (sequential exposure). Stars indicate observed values, violins indicate exposures among permuted random transmission clusters. Spatiotemporal exposure is enriched in transmission clusters compared to permuted groups of patients of the same size and patient makeup (admission positive and acquisition patients) as the observed clusters (permutation tests, p<0.001 for all locations). Sequential exposure is not enriched in transmission clusters compared to random clusters (permutation tests, p>0.60 for all locations.)

Lastly, we sought to examine transmission clusters more holistically to gain insight into generalizable principles regarding KPC-Kp transmission pathways in the facility. Visual inspection of transmission clusters revealed several themes that manifested across multiple clusters (**Figure 7**). These themes included: i) transmission between cohort and non-cohort locations, presumably due to movement of a common healthcare worker between locations, or use of common location by a KPC-Kp positive and KPC-Kp negative patients (e.g., physical therapy gym) (**Figure 7A)**, ii) lapses in cohorting e.g. transmission due to housing a known positive patient in the same location as a negative patient (**Figure 7B)**, iii) apparent false-negative surveillance cultures allowing clusters to propagate undetected (**Figure 7C**), iv) undetected patient or environmental sources as evidenced by temporal gaps in clusters (**Figure 7D**) and v) likely exposure between cluster patients that occurred in an outside facility prior to admission (**Figure 7E**). The plausible routes of transmission illustrated in these vignettes are not mutually exclusive of one another and evidence supporting multiple routes occurred in several clusters.

**Figure 7:**
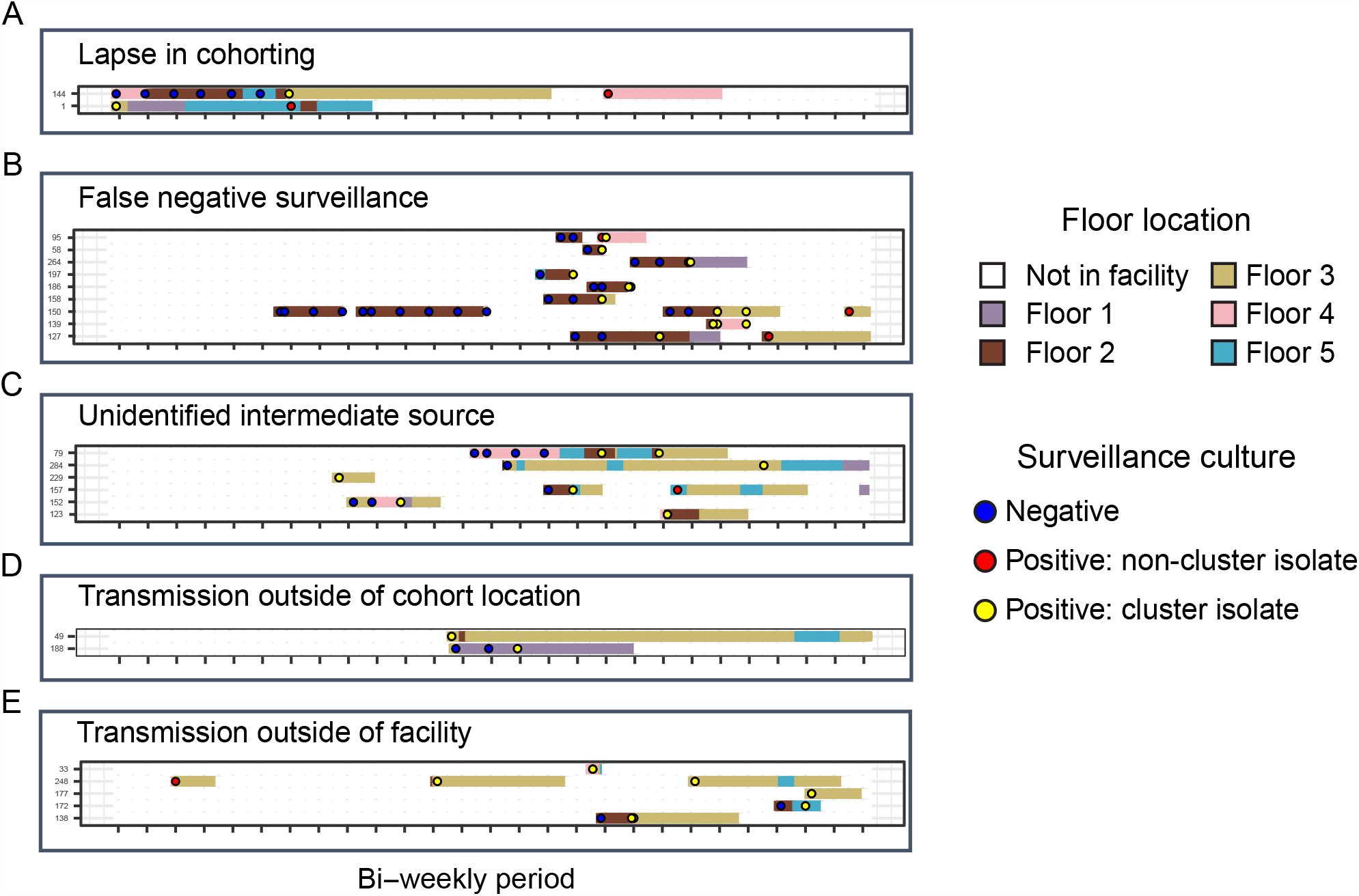
Descriptive vignettes from transmission clusters detected through the integration of genomic and surveillance data illustrate putative routes of uncontrolled transmission. Patients are indicated on the y-axis and time is on the x-axis. Putative route of transmission within each cluster is indicated in the text above the cluster. Surveillance culturing information is indicated by the circles, and floor location in the LTACH is indicated by the colored rectangles. **A**. Transmission between positive admission positive patient 1 to acquisition patient 144. Both were on the teal ward while patient 1 was positive and patient 144 was negative. **B**. No admission positive patient precedes several acquisition patients in this cluster, therefore false negative surveillance of a patient in the cluster or a patient not captured in the study is likely source. **C**. Lack of spatiotemporal exposures between several patients indicate missing intermediate source patient undetected by surveillance culturing. **D**. Transmission between two patients who did not reside on the same ward indicates potential escape from cohort location, or transmission at a common location, or via an unidentified common healthcare worker source in the facility. **E**. Multiple admission positive patients and lack of spatiotemporal exposures in the LTACH indicates potential transmission outside of the facility before admission to the LTACH.

## Discussion

Whole-genome sequencing has become the gold-standard for studying the spread of infections in healthcare settings^32^. However, harnessing WGS to optimize infection prevention is hindered by the lack of standardized criteria to detect transmission occurring within a healthcare facility, particularly for widespread, epidemic strains that are continuously imported into healthcare facilities in endemic settings^14,33^. Here, we sought to dissect intra-facility transmission pathways in a high prevalence, endemic setting by leveraging a comprehensive sample collection in which importation and acquisition events were comprehensively discerned via whole-hospital admission and biweekly KPC-Kp surveillance culturing for a one-year period. This comprehensive sampling allowed us to demonstrate the pitfalls associated with imposing an SNV threshold to delineate transmission, and then to apply a threshold-free approach to group acquisition events into transmission clusters traced back to importation events. Examination of these threshold-free transmission clusters yielded insight into the genetic diversity underlying true transmission pairs and allowed for deconvolution of transmission pathways in a setting with extremely high colonization prevalence.

Our unbiased view of the genetic diversity underlying putative transmission events clearly demonstrated how the imposition of a strict SNV threshold could lead to significant false positive and false negative transmission inferences. The primary source of false-negative inferences is the accumulation of genetic variation during prolonged asymptomatic colonization, which can manifest in large, and uneven, genetic distances between patients linked by transmission within the facility^12,15,16^. Moreover, we were able to show that this intra-patient diversity can be further amplified by the emergence of hypermutator strains, which yield genetic distances that would elude any approach grounded in SNV thresholds^11,13^. Conversely, SNV threshold approaches can also lead to false positive transmission inferences due to the importation of closely related strains that may be linked by recent transmission at a connected healthcare facility^11,14,34,35^. This dependence on local connectivity of regional healthcare networks and the transmission dynamics at these connected healthcare facilities is further reason to be skeptical of the potential for a one-size fits all SNV threshold that can be broadly applied in different contexts. The success of our approach, even in a high prevalence endemic setting, indicates that approaches grounded in genetic context may be more robust than those relying on genetic distance.

Our study has several limitations related to biases in sampling. First, although we sampled 94% of patients in the facility during the yearlong study, only a single or small number of colonies (representative unique morphologies) were collected and sequenced per patient, and patients were not re-surveilled systematically once identified to carry KPC-Kp. Therefore, we may have missed cases where a patient imported multiple strains into the facility, or where a patient acquired a second KPC-Kp strain later in their hospital stay. Either of these limitations could potentially account for some of the cases where we were unable to identify a cluster source patient. Second, our lack of knowledge of where patients were prior to admission prevents us from understanding how transmission at connected healthcare facilities influenced grouping of patients in clusters. We hypothesize that these transmission events outside of the facility accounted for cases where admission-positive patients were not the first members of their cluster to test positive. Third, there is an inherent limit of detection of surveillance culturing, which is likely associated with variation in the density of KPC-Kp colonization in the gut^36^. Thus, false-negative surveillance cultures could have resulted in patients who imported KPC-Kp either being inferred to have acquired it in the facility, or remaining undetected over the course of the study. These cryptic cases could account for clusters lacking admission positive patients and/or be the cause of clusters where not all transmission could be explained by spatiotemporal exposures between patients. Fourth, we studied patients in a single LTACH with high prevalence of KPC-Kp. Results may not be generalizable to lower prevalence settings.

Overall, our results highlight the potential for WGS to improve infection surveillance and prevention when combined with appropriate sampling and analytical strategies that are jointly tailored to generate actionable hypotheses. The SNV threshold-free approach applied here could be deployed with only admission and discharge surveillance culturing, although higher resolution sampling would facilitate more precise delineation of transmission pathways within clusters.

Importantly, by relying on shared variants, as opposed to genetic distances, inferences should be agnostic to the specific MDRO species or strain, thereby circumventing the need for constant refinement of discriminatory criteria and facilitating clearer interpretation and more effective intervention by healthcare epidemiologists.

## Data Availability

Upon acceptance into a peer reviewed journal, whole genome sequencing data will be available publicly under NCBI BioProject PRJNA603790.

## Acknowledgments

We thank the patients and staff of the long-term acute-care hospital (LTACH) for their gracious participation in this study; Ali Pirani for bioinformatics support and members of the Snitkin lab and the Rush University/University of Michigan genomics working group for critical review of the manuscript.

## Supplementary materials

**Figure S1.**
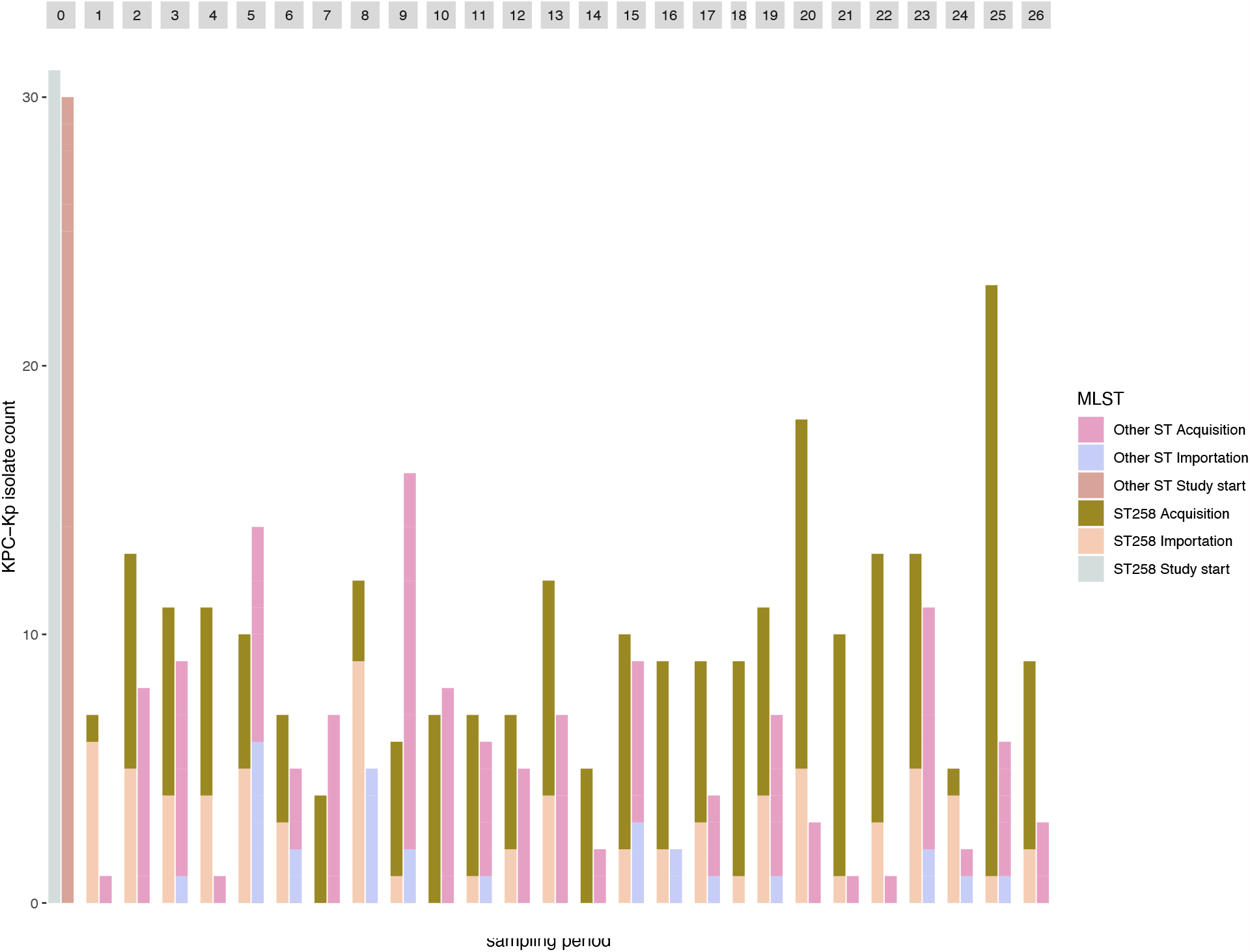
Endemicity of KPC-Kp in the LTACH is due to extensive importation and acquisition of multiple KPC-Kp strains throughout the study. MLST of isolates obtained through bi-weekly rectal surveillance culturing of LTACH patients. Grey boxes indicate the study start (time 0) and 14-day surveillance periods throughout the study. Bars indicate the KPC-Kp isolates collected at the beginning of the study, after importation or acquisition (>3 days after a patient was in the LTACH). Legend indicates ST258 or other MLST of isolates.

**Figure S2.**
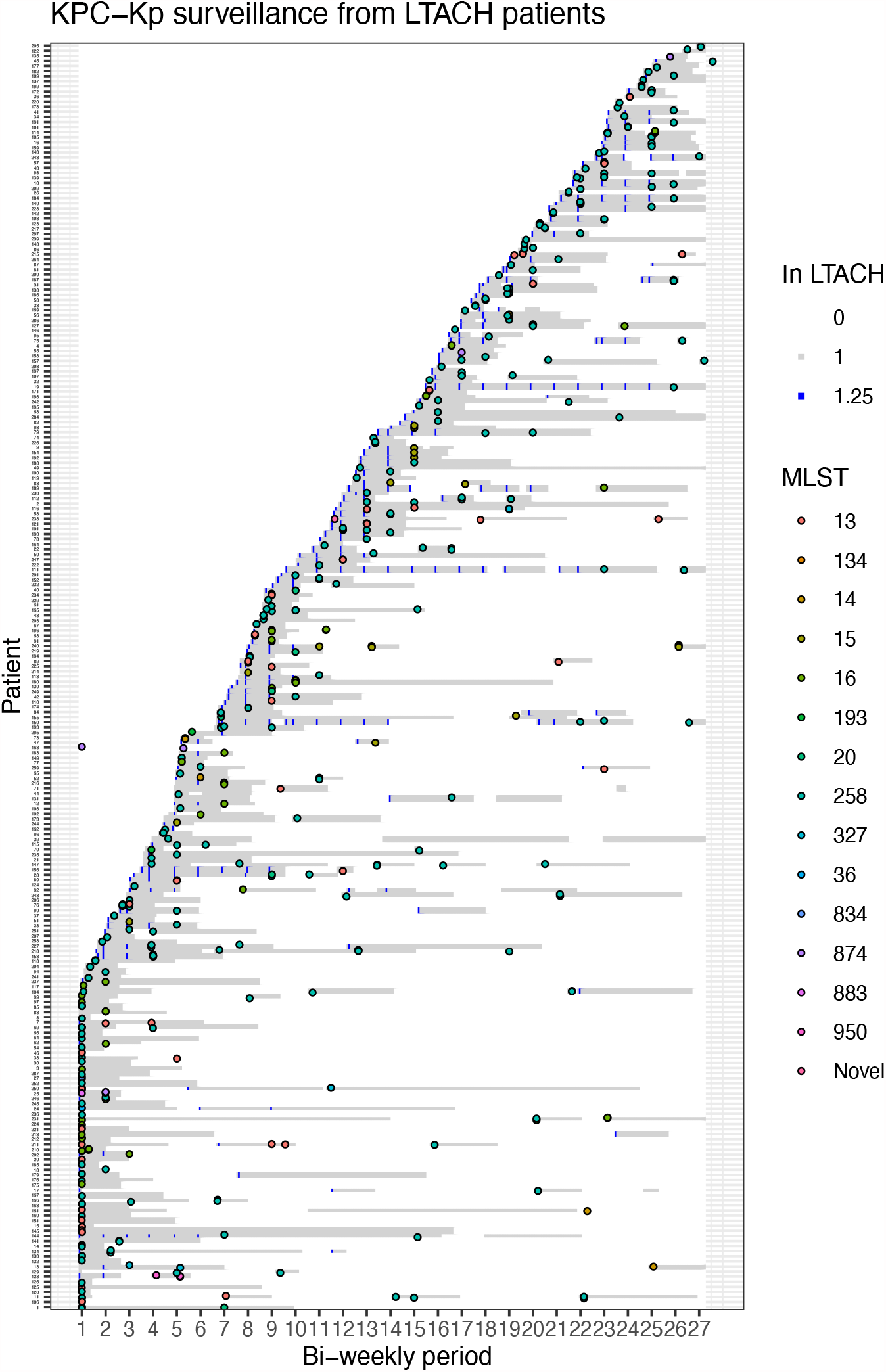
Complexity of discerning KPC-Kp transmission chains in the LTACH is illustrated by extensive importation and acquisition of multiple strains and patients with shared time in the LTACH. **A**. Patient bed trace showing surveillance of patients who either imported or acquired KPC-Kp colonization in the LTACH. The order of patients on the Y-axis is by first date in the LTACH. Grey bars indicate patients in the LTACH, white indicates outside of the facility, colored circles indicate the MLSTs of isolates obtained by positive surveillance cultures from a patient on a collection date (x axis). **B-G**. Subset trace plots for patients with indicated MLSTs with plausible in-LTACH transmission links indicated in **Table 1**.

**Figure S3:**
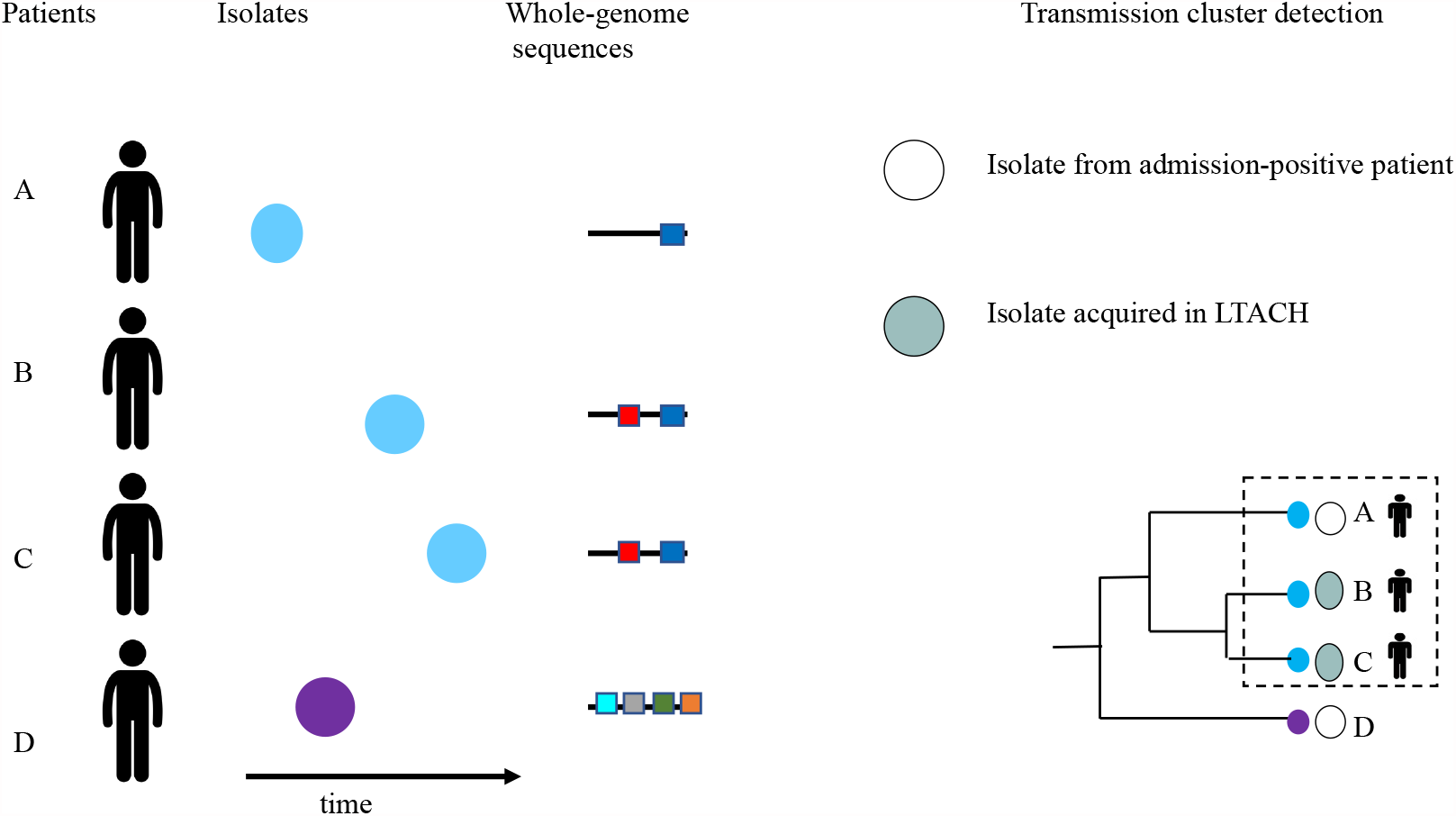
Schematic of genomic transmission cluster detection strategy that integrates shared variants from whole-genome sequences with surveillance data. Shared variants in whole genome sequences (black lines, variants are colored boxes) from isolates sampled from patients are used to construct a maximum parsimony phylogeny. Transmission clusters are defined by the maximum subtree in the phylogeny that contains isolates from a single admission-positive patient who imported the isolate from outside the facility. Valid transmission clusters must contain at least a single unique variant (yellow box) that distinguishes cluster from non-cluster isolates (A, B and C isolates (dashed box) vs isolate D), and at least two patients including at least one acquisition patient who acquired KPC-Kp colonization in the LTACH. Clusters with multiple admission positive/study-start patients are valid if the isolates share unique variants with other cluster members. Clusters with no admission positive patients are valid if there existed no subtree that contained an admission positive isolate.

**Figure S4: Patient bed traces and surveillance information for the 49 transmission clusters detected through the integration of genomic and surveillance data**. Patients are indicated on the Y axis and time is on the X -axis. **See PDF: all_cluster_trace_plots.pdf**

## References

1. Magill, S. S. et al. Changes in Prevalence of Health Care–Associated Infections in U.S. Hospitals. N. Engl. J. Med. 379, 1732–1744 (2018).

2. Healthcare-associated Infections (HAI) Progress Report | HAI | CDC. http://www.cdc.gov/hai/surveillance/progress-report/index.html.

3. Snitkin, E. S. et al. Tracking a Hospital Outbreak of Carbapenem-Resistant Klebsiella pneumoniae with Whole-Genome Sequencing. Sci. Transl. Med. 4, 148ra116–148ra116 (2012).

4. Köser, C. U. et al. Rapid Whole-Genome Sequencing for Investigation of a Neonatal MRSA Outbreak. N. Engl. J. Med. 366, 2267–2275 (2012).

5. Stoesser, N. et al. Dynamics of MDR Enterobacter cloacae outbreaks in a neonatal unit in Nepal: insights using wider sampling frames and next-generation sequencing. J. Antimicrob. Chemother. 70, 1008–1015 (2015).

6. Sherry, N. L. et al. Pilot study of a combined genomic and epidemiologic surveillance program for hospital-acquired multidrug-resistant pathogens across multiple hospital networks in Australia. Infect. Control Hosp. Epidemiol. 42, 573–581 (2021).

7. Gouliouris, T. et al. Quantifying acquisition and transmission of Enterococcus faecium using genomic surveillance. Nat. Microbiol. 6, 103–111 (2021).

8. Coll, F. et al. Definition of a genetic relatedness cutoff to exclude recent transmission of meticillin-resistant Staphylococcus aureus: a genomic epidemiology analysis. Lancet Microbe 1, e328–e335 (2020).

9. Tosas Auguet, O. et al. Frequent Undetected Ward-Based Methicillin-Resistant Staphylococcus aureus Transmission Linked to Patient Sharing Between Hospitals. Clin. Infect. Dis. Off. Publ. Infect. Dis. Soc. Am. 66, 840–848 (2018).

10. Stimson, J. et al. Beyond the SNP Threshold: Identifying Outbreak Clusters Using Inferred Transmissions. Mol. Biol. Evol. 36, 587–603 (2019).

11. Han, J. H. et al. Whole-Genome Sequencing To Identify Drivers of Carbapenem-Resistant Klebsiella pneumoniae Transmission within and between Regional Long-Term Acute-Care Hospitals. Antimicrob. Agents Chemother. 63, (2019).

12. Hall, M. D. et al. Improved characterisation of MRSA transmission using within-host bacterial sequence diversity. eLife 8, (2019).

13. David, S. et al. Epidemic of carbapenem-resistant Klebsiella pneumoniae in Europe is driven by nosocomial spread. Nat. Microbiol. 4, 1919–1929 (2019).

14. Wang, J. et al. Application of combined genomic and transfer analyses to identify factors mediating regional spread of antibiotic resistant bacterial lineages. Clin. Infect. Dis. Off. Publ. Infect. Dis. Soc. Am. (2020) doi:10.1093/cid/ciaa364.

15. Didelot, X., Walker, A. S., Peto, T. E., Crook, D. W. & Wilson, D. J. Within-host evolution of bacterial pathogens. Nat. Rev. Microbiol. 14, 150–162 (2016).

16. Paterson, G. K. et al. Capturing the cloud of diversity reveals complexity and heterogeneity of MRSA carriage, infection and transmission. Nat. Commun. 6, 6560 (2015).

17. Haverkate, M. R. et al. Duration of Colonization With Klebsiella pneumoniae Carbapenemase-Producing Bacteria at Long-Term Acute Care Hospitals in Chicago, Illinois. Open Forum Infect. Dis. 3, (2016).

18. O’Fallon, E., Gautam, S. & D’Agata, E. M. C. Colonization with multidrug-resistant gram-negative bacteria: prolonged duration and frequent cocolonization. Clin. Infect. Dis. Off. Publ. Infect. Dis. Soc. Am. 48, 1375–1381 (2009).

19. Golubchik, T. et al. Within-host evolution of Staphylococcus aureus during asymptomatic carriage. PloS One 8, e61319 (2013).

20. Hayden, M. K. et al. Prevention of Colonization and Infection by Klebsiella pneumoniae Carbapenemase–Producing Enterobacteriaceae in Long-term Acute-Care Hospitals. Clin. Infect. Dis. 60, 1153–1161 (2015).

21. Lolans, K., Calvert, K., Won, S., Clark, J. & Hayden, M. K. Direct Ertapenem Disk Screening Method for Identification of KPC-Producing Klebsiella pneumoniae and Escherichia coli in Surveillance Swab Specimens. J. Clin. Microbiol. 48, 836–841 (2010).

22. Haverkate, M. R. et al. Modeling Spread of KPC-Producing Bacteria in Long-Term Acute Care Hospitals in the Chicago Region, USA. Infect. Control Hosp. Epidemiol. 36, 1148– 1154 (2015).

23. Babraham Bioinformatics - FastQC A Quality Control tool for High Throughput Sequence Data. https://www.bioinformatics.babraham.ac.uk/projects/fastqc/.

24. Bolger, A. M., Lohse, M. & Usadel, B. Trimmomatic: a flexible trimmer for Illumina sequence data. Bioinformatics 30, 2114–2120 (2014).

25. Hawken, S. E. et al. Cohorting KPC+ Klebsiella pneumoniae (KPC-Kp)-positive patients: A genomic exposé of cross-colonization hazards in a long-term acute-care hospital (LTACH). Infect. Control Hosp. Epidemiol. 41, 1162–1168 (2020).

26. Han, J. H. et al. Whole-Genome Sequencing To Identify Drivers of Carbapenem-Resistant Klebsiella pneumoniae Transmission within and between Regional Long-Term Acute-Care Hospitals. Antimicrob. Agents Chemother. 63, e01622–19 (2019).

27. Li, H. et al. The Sequence Alignment/Map format and SAMtools. Bioinformatics 25, 2078– 2079 (2009).

28. Li, H. & Durbin, R. Fast and accurate short read alignment with Burrows–Wheeler transform. Bioinformatics 25, 1754–1760 (2009).

29. Treepong, P. et al. panISa: ab initio detection of insertion sequences in bacterial genomes from short read sequence data. Bioinformatics 34, 3795–3800 (2018).

30. Bowers, J. R. et al. Genomic Analysis of the Emergence and Rapid Global Dissemination of the Clonal Group 258 Klebsiella pneumoniae Pandemic. PloS One 10, e0133727 (2015).

31. Couce, A. et al. Mutator genomes decay, despite sustained fitness gains, in a long-term experiment with bacteria. Proc. Natl. Acad. Sci. 114, E9026–E9035 (2017).

32. Gerner-Smidt, P. et al. Whole Genome Sequencing: Bridging One-Health Surveillance of Foodborne Diseases. Front. Public Health 7, (2019).

33. Sherry, N. L. et al. Pilot study of a combined genomic and epidemiologic surveillance program for hospital-acquired multidrug-resistant pathogens across multiple hospital networks in Australia. Infect. Control Hosp. Epidemiol. 1–9 (undefined/ed) doi:10.1017/ice.2020.1253.

34. Donker, T. et al. Population genetic structuring of methicillin-resistant Staphylococcus aureus clone EMRSA-15 within UK reflects patient referral patterns. Microb. Genomics 3, e000113 (2017).

35. Chang, H.-H. et al. Identifying the effect of patient sharing on between-hospital genetic differentiation of methicillin-resistant Staphylococcus aureus. Genome Med. 8, 18 (2016).

36. Haverkate, M. R. et al. Modeling spread of KPC-producing bacteria in long-term acute care hospitals in the Chicago region, USA. Infect. Control Hosp. Epidemiol. 36, 1148–1154 (2015).

