## Supplemental_Figure_4 for "Threshold-free genomic cluster detection to track transmission pathways in healthcare settings"

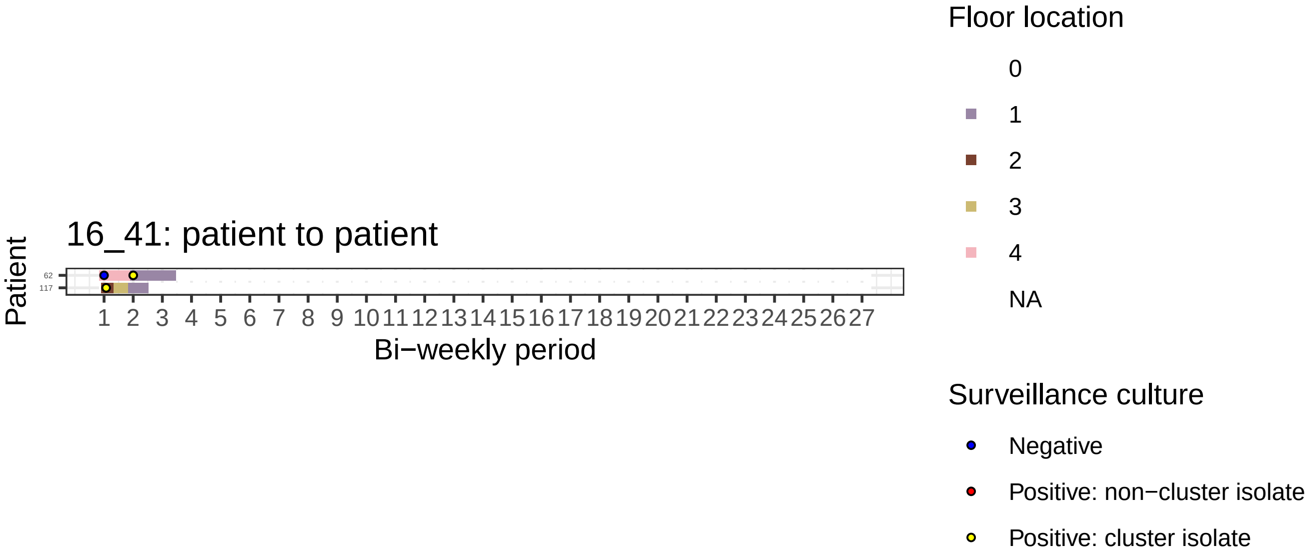

16_41: patient to patient
Patient
62
117
1 2 3 4 5 6 7 8 9 10 11 12 13 14 15 16 17 18 19 20 21 22 23 24 25 26 27
Bi−weekly period
Floor location
0
1
2
3
4
NA
Surveillance culture
Negative
Positive: non−cluster isolate
Positive: cluster isolate

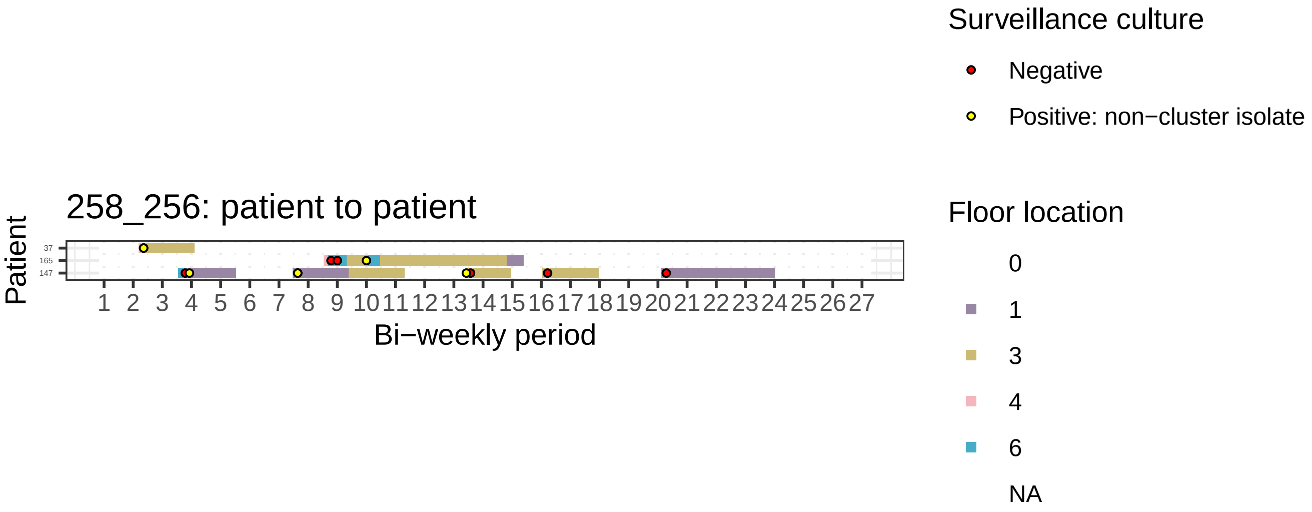

258_256: patient to patient
Patient
37
165
147
Bi−weekly period
Surveillance culture
Negative
Positive: non−cluster isolate
Floor location
0
1
3
4
6
NA

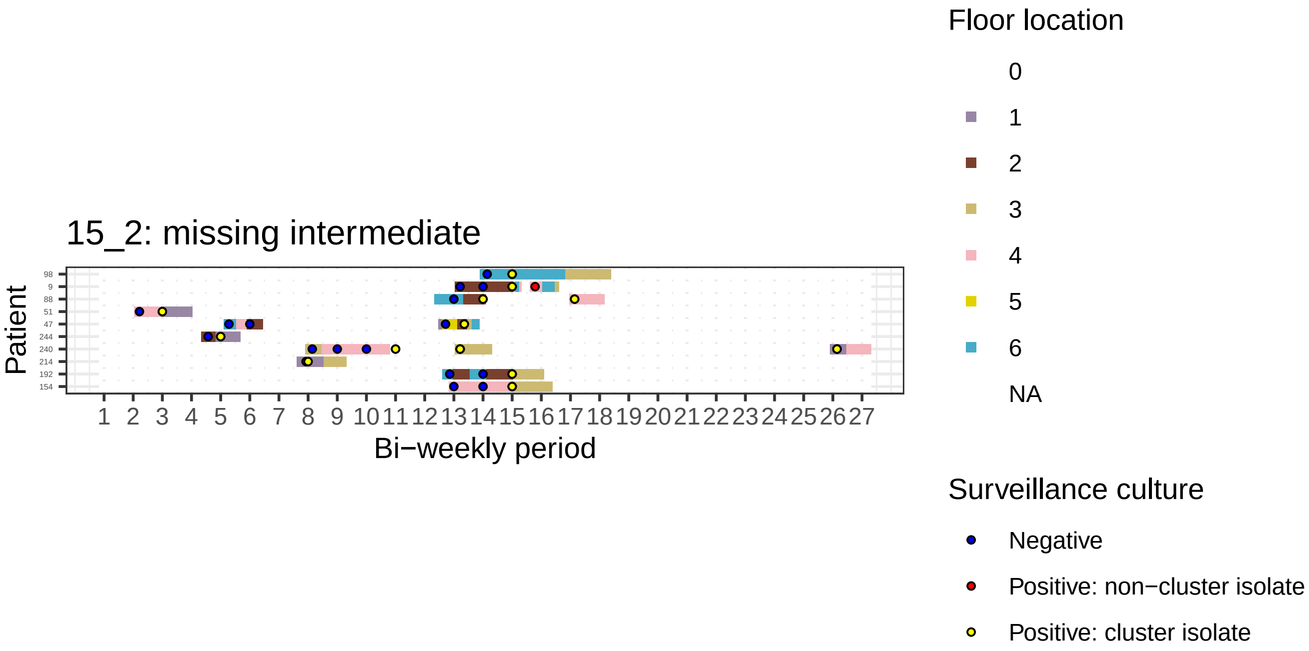

15_2: missing intermediate
Patient
98
9
88
51
47
244
240
214
192
154
1 2 3 4 5 6 7 8 9 10 11 12 13 14 15 16 17 18 19 20 21 22 23 24 25 26 27
Bi−weekly period
Floor location
0
1
2
3
4
5
6
NA
Surveillance culture
Negative
Positive: non−cluster isolate
Positive: cluster isolate

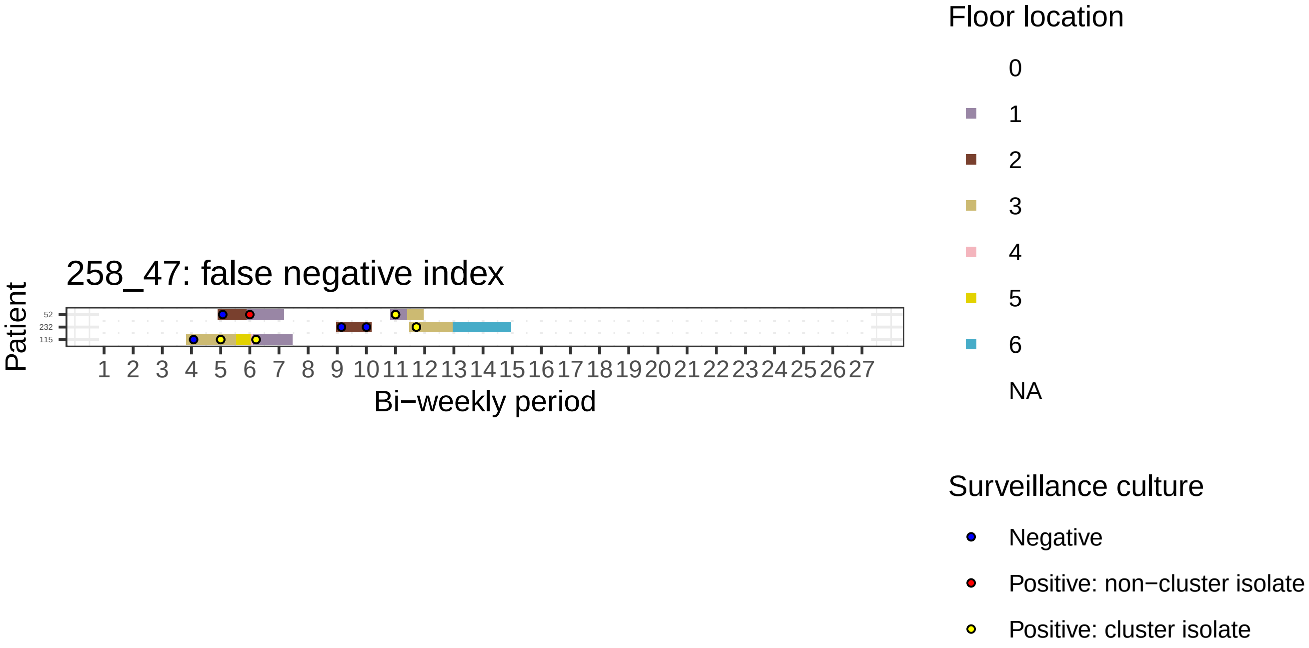

258_47: false negative index
Patient
52
232
115
1 2 3 4 5 6 7 8 9 10 11 12 13 14 15 16 17 18 19 20 21 22 23 24 25 26 27
Bi−weekly period
Floor location
0
1
2
3
4
5
6
NA
Surveillance culture
Negative
Positive: non−cluster isolate
Positive: cluster isolate

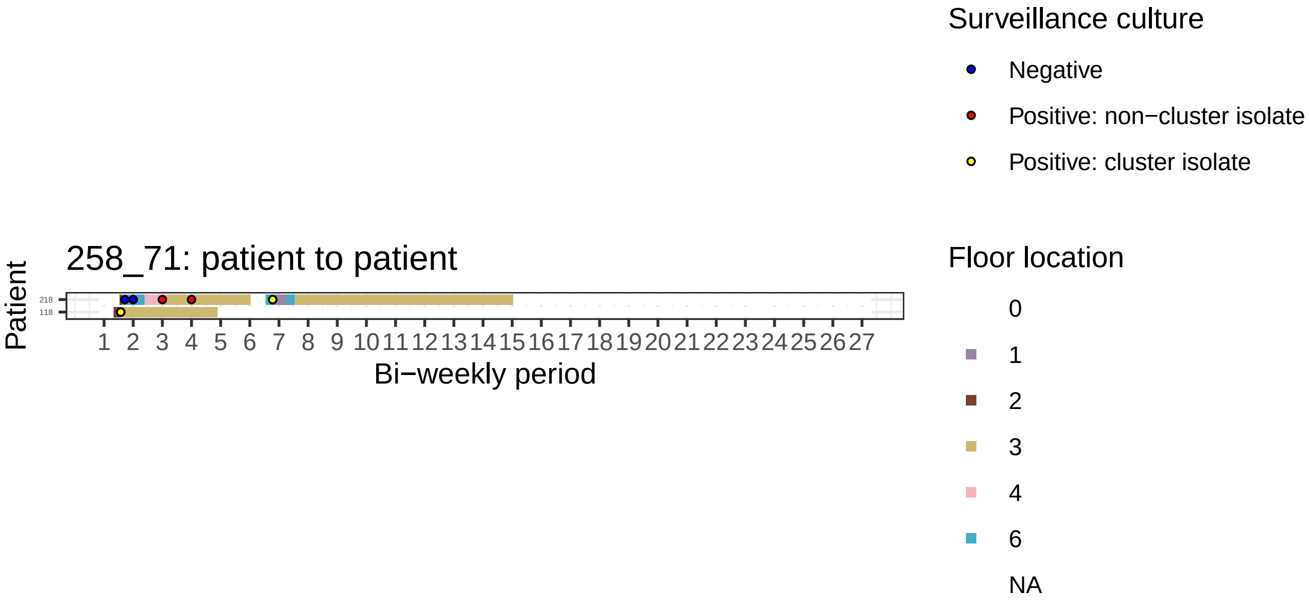
258_71: patient to patient
Patient
218
118
1 2 3 4 5 6 7 8 9 10 11 12 13 14 15 16 17 18 19 20 21 22 23 24 25 26 27
Bi−weekly period
Surveillance culture
Negative
Positive: non−cluster isolate
Positive: cluster isolate
Floor location
0
1
2
3
4
6
NA

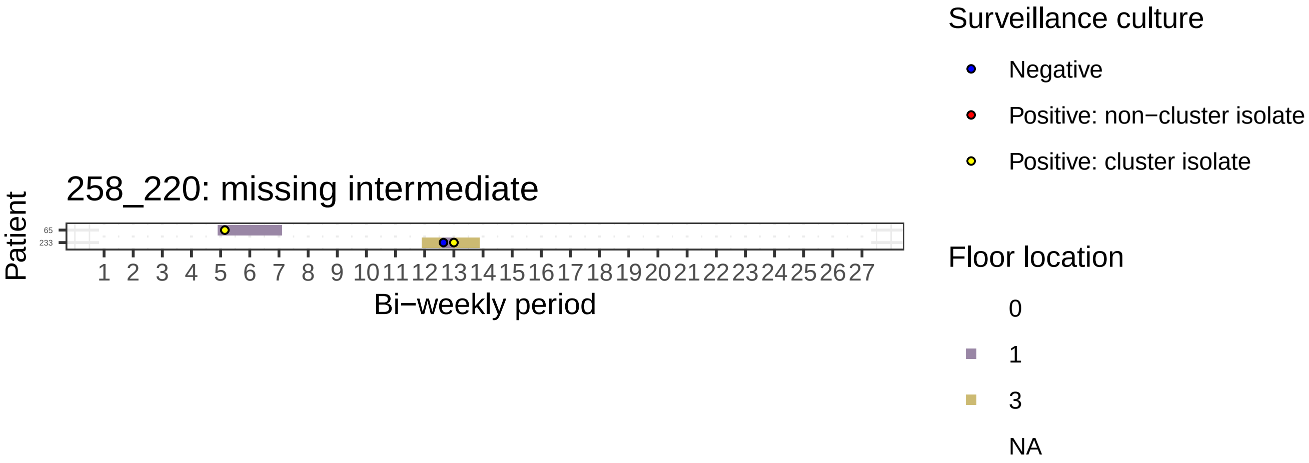
258_220: missing intermediate
Patient
65
233
Bi−weekly period
Surveillance culture
Negative
Positive: non−cluster isolate
Positive: cluster isolate
Floor location
0
1
3
NA

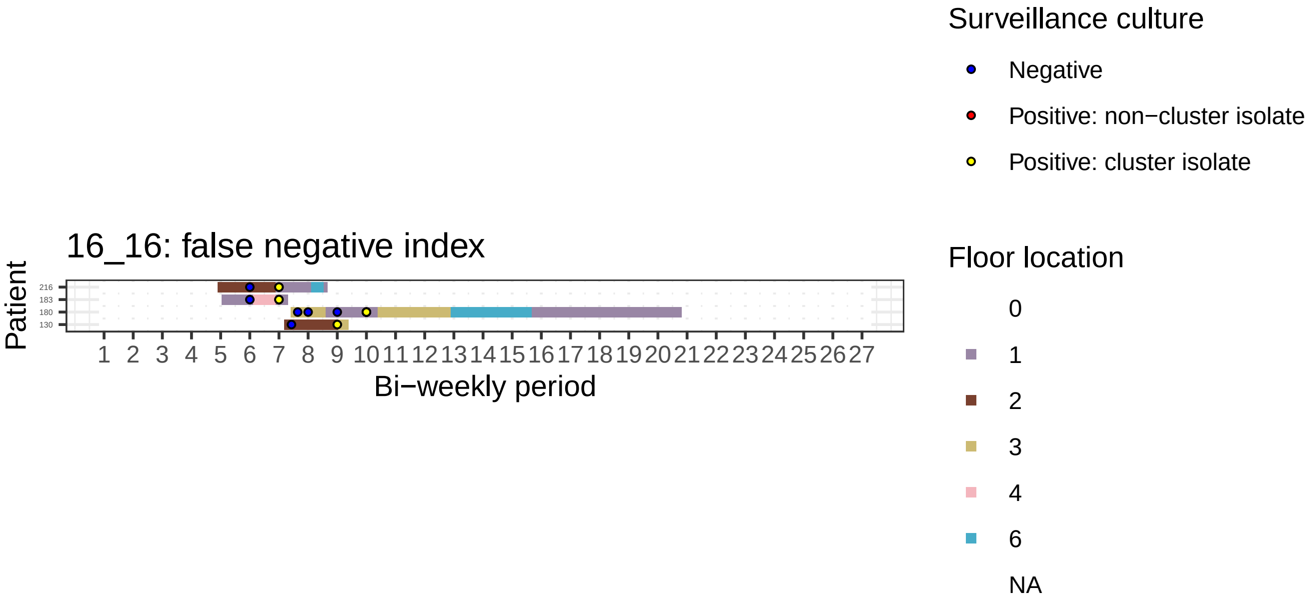
16_16: false negative index
Patient
216
183
180
130
1 2 3 4 5 6 7 8 9 10 11 12 13 14 15 16 17 18 19 20 21 22 23 24 25 26 27
Bi−weekly period
Surveillance culture
Negative
Positive: non−cluster isolate
Positive: cluster isolate
Floor location
0
1
2
3
4
6
NA

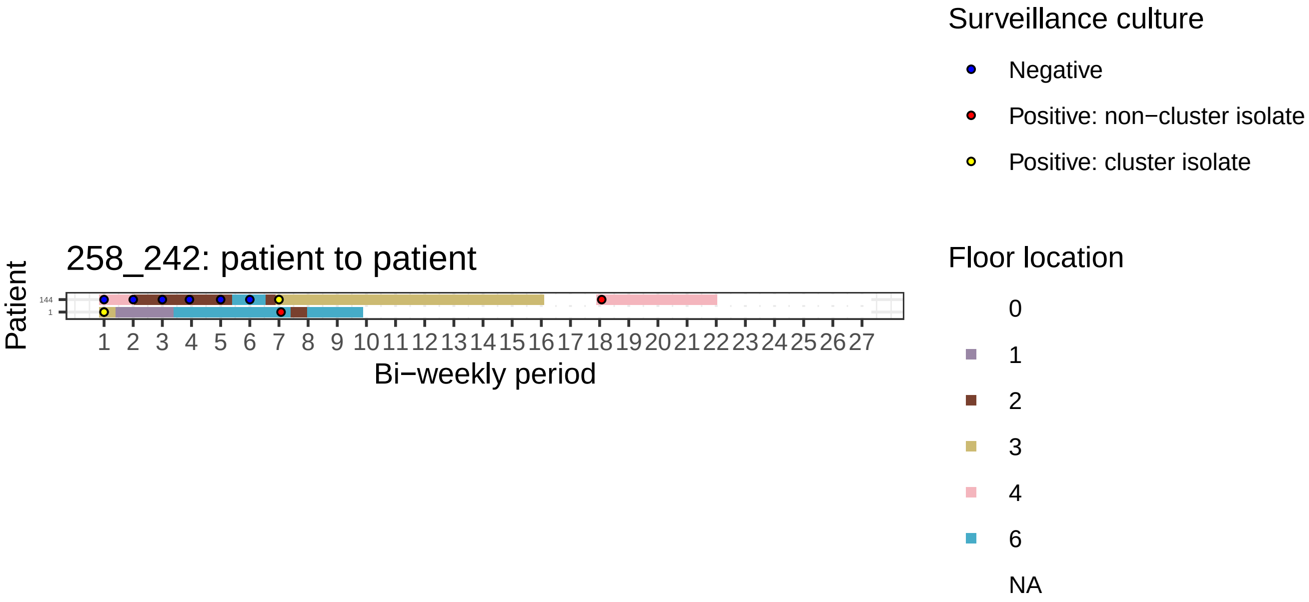

258_242: patient to patient
Patient
144
1
Bi−weekly period
Surveillance culture
Negative
Positive: non−cluster isolate
Positive: cluster isolate
Floor location
0
1
2
3
4
6
NA

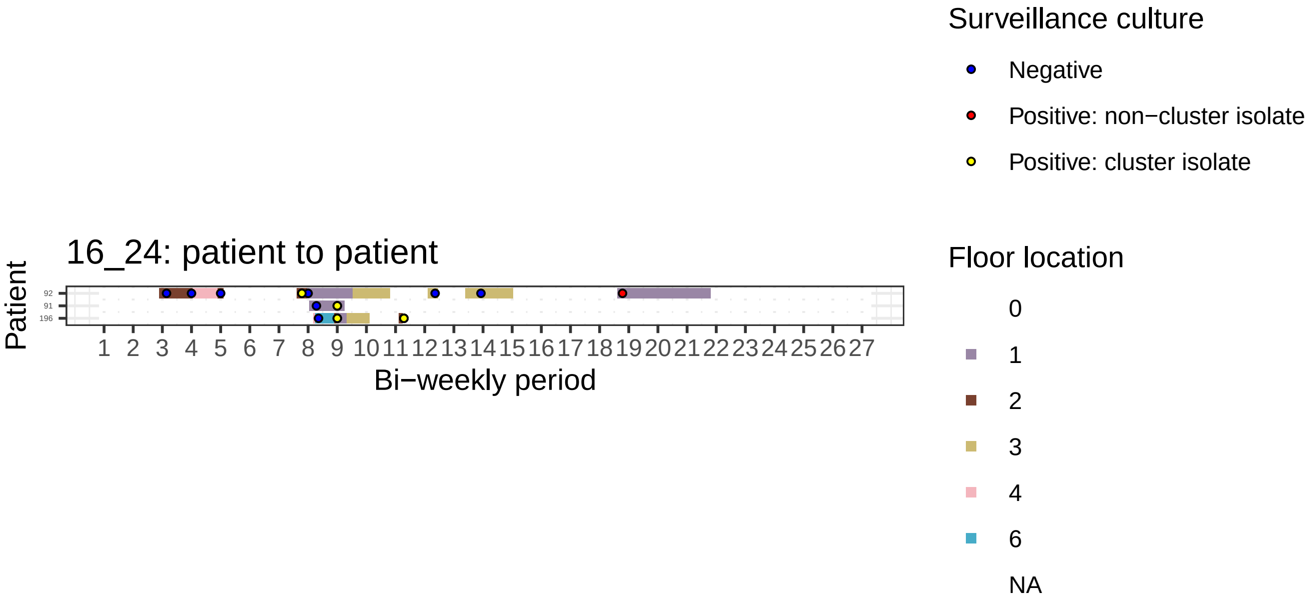
16_24: patient to patient
Patient
92
91
196
1 2 3 4 5 6 7 8 9 10 11 12 13 14 15 16 17 18 19 20 21 22 23 24 25 26 27
Bi−weekly period
Surveillance culture
Negative
Positive: non−cluster isolate
Positive: cluster isolate
Floor location
0
1
2
3
4
6
NA

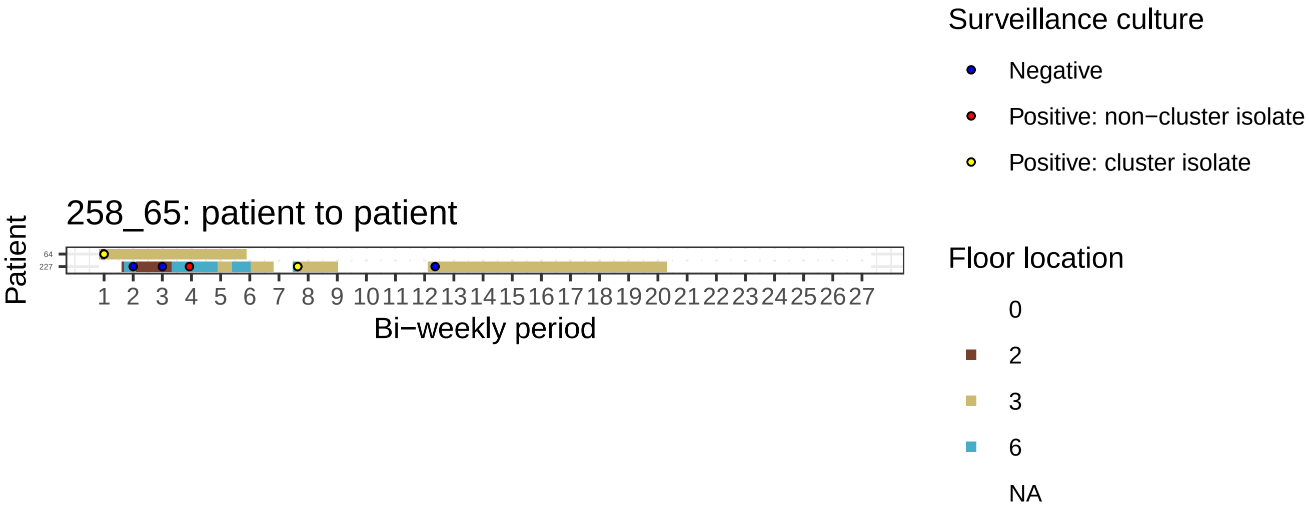
258_65: patient to patient
Patient
64
227
1 2 3 4 5 6 7 8 9 10 11 12 13 14 15 16 17 18 19 20 21 22 23 24 25 26 27
Bi−weekly period
Surveillance culture
Negative
Positive: non−cluster isolate
Positive: cluster isolate
Floor location
0
2
3
6
NA

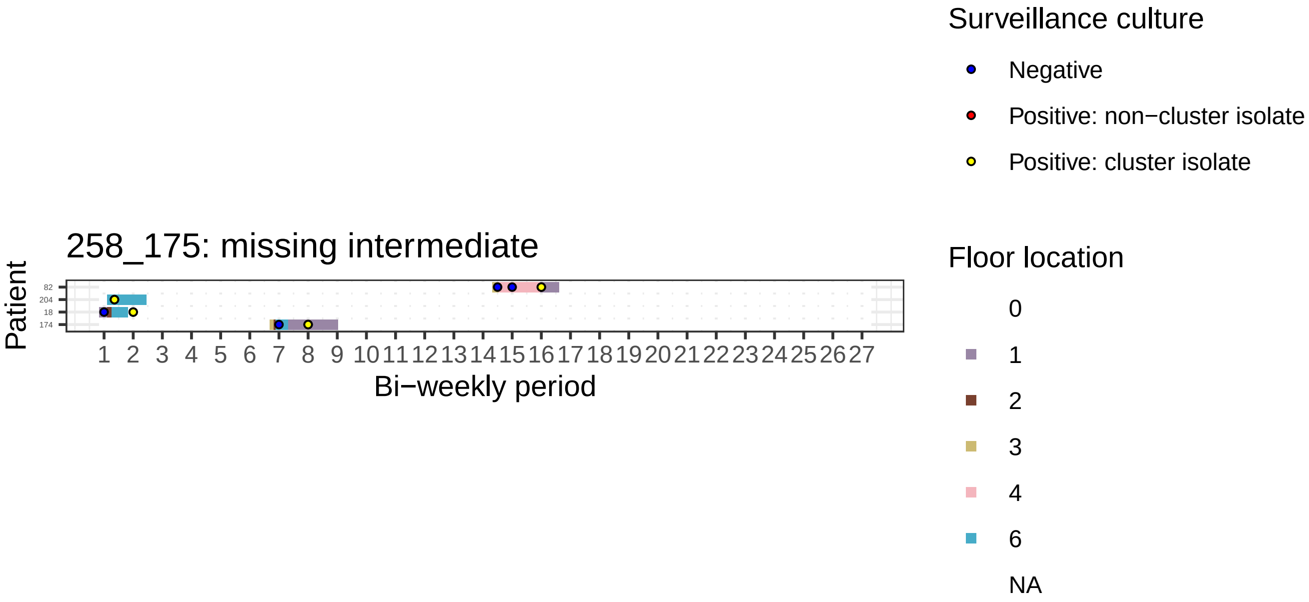

258_175: missing intermediate
Patient
82
204
18
174
1 2 3 4 5 6 7 8 9 10 11 12 13 14 15 16 17 18 19 20 21 22 23 24 25 26 27
Bi−weekly period
Surveillance culture
Negative
Positive: non−cluster isolate
Positive: cluster isolate
Floor location
0
1
2
3
4
6
NA

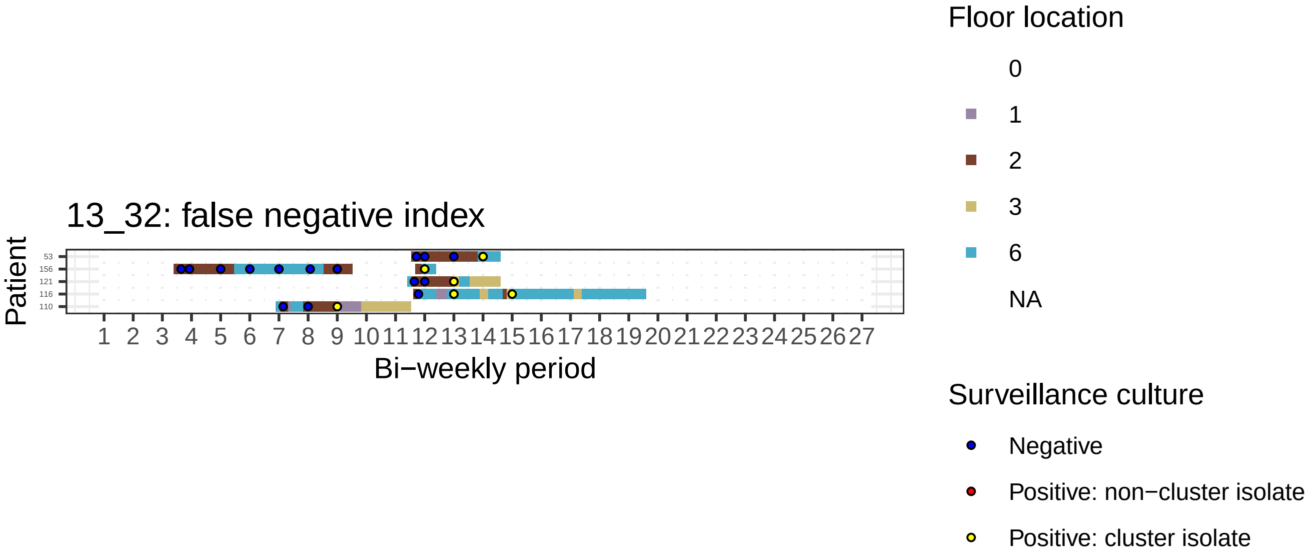
13_32: false negative index
Patient
53
156
121
116
110
1 2 3 4 5 6 7 8 9 10 11 12 13 14 15 16 17 18 19 20 21 22 23 24 25 26 27
Bi−weekly period
Floor location
0
1
2
3
6
NA
Surveillance culture
Negative
Positive: non−cluster isolate
Positive: cluster isolate

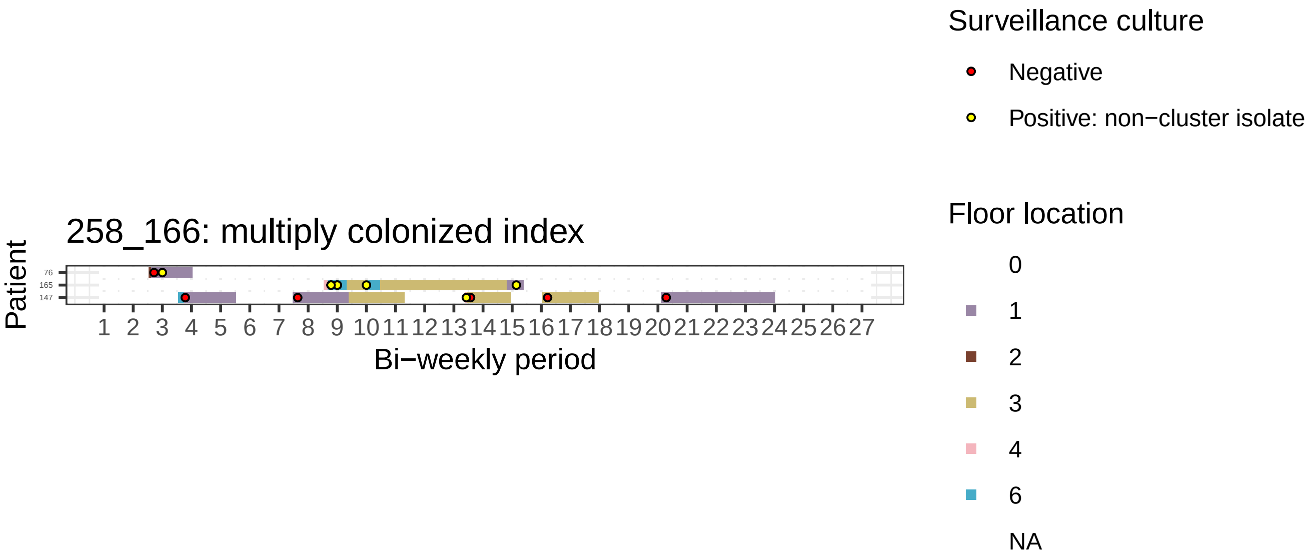
258_166: multiply colonized index
Patient
76
165
147
1 2 3 4 5 6 7 8 9 10 11 12 13 14 15 16 17 18 19 20 21 22 23 24 25 26 27
Bi−weekly period
Surveillance culture
Negative
Positive: non−cluster isolate
Floor location
0
1
2
3
4
6
NA

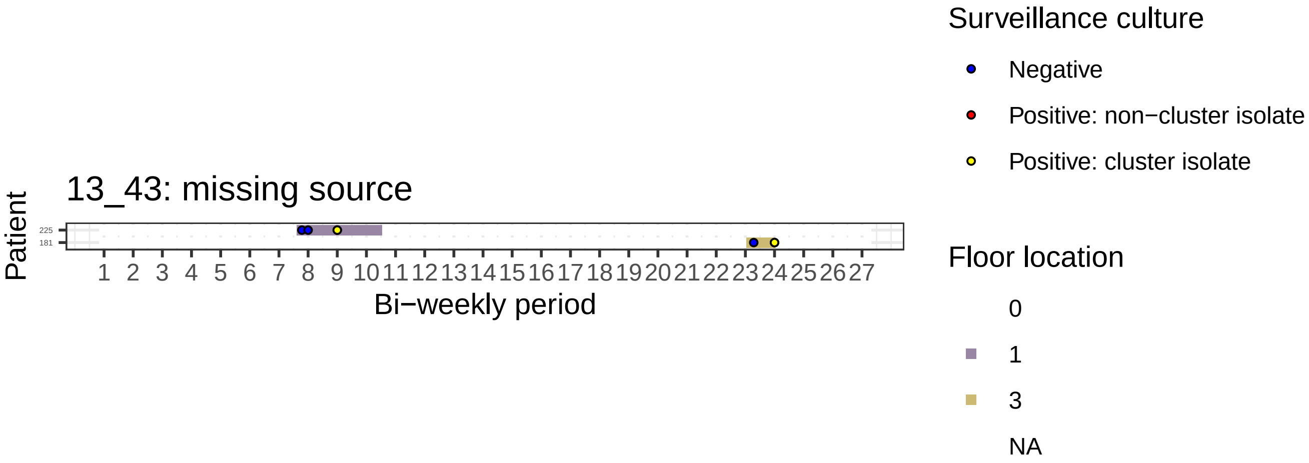

13_43: missing source
Patient
225
181
Bi−weekly period
Surveillance culture
Negative
Positive: non−cluster isolate
Positive: cluster isolate
Floor location
0
1
3
NA

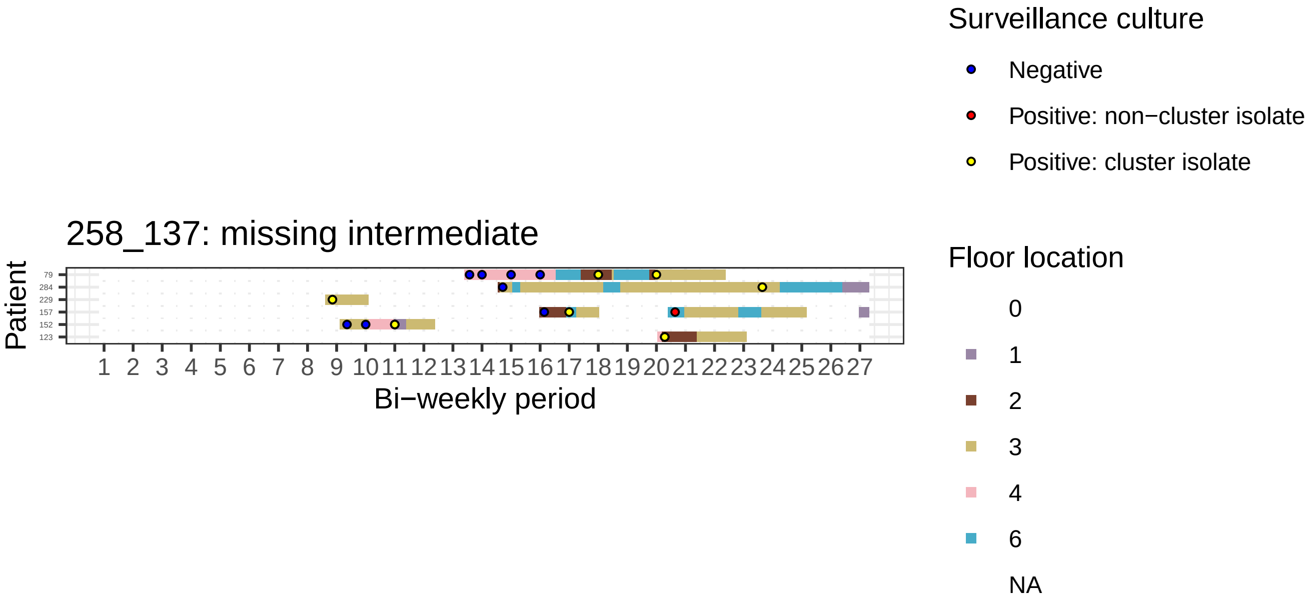

258_137: missing intermediate
Patient
79
284
229
157
152
123
1 2 3 4 5 6 7 8 9 10 11 12 13 14 15 16 17 18 19 20 21 22 23 24 25 26 27
Bi−weekly period
Surveillance culture
Negative
Positive: non−cluster isolate
Positive: cluster isolate
Floor location
0
1
2
3
4
6
NA

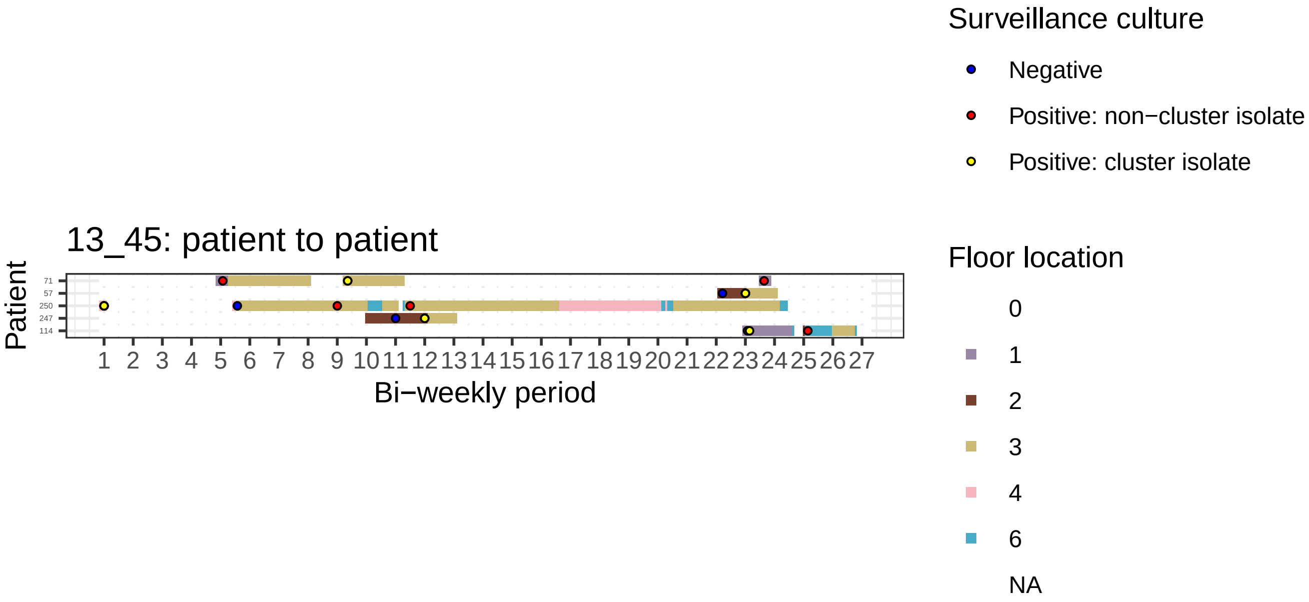

13_45: patient to patient
Patient
71
57
250
247
114
1 2 3 4 5 6 7 8 9 10 11 12 13 14 15 16 17 18 19 20 21 22 23 24 25 26 27
Bi−weekly period
Surveillance culture
Negative
Positive: non−cluster isolate
Positive: cluster isolate
Floor location
0
1
2
3
4
6
NA

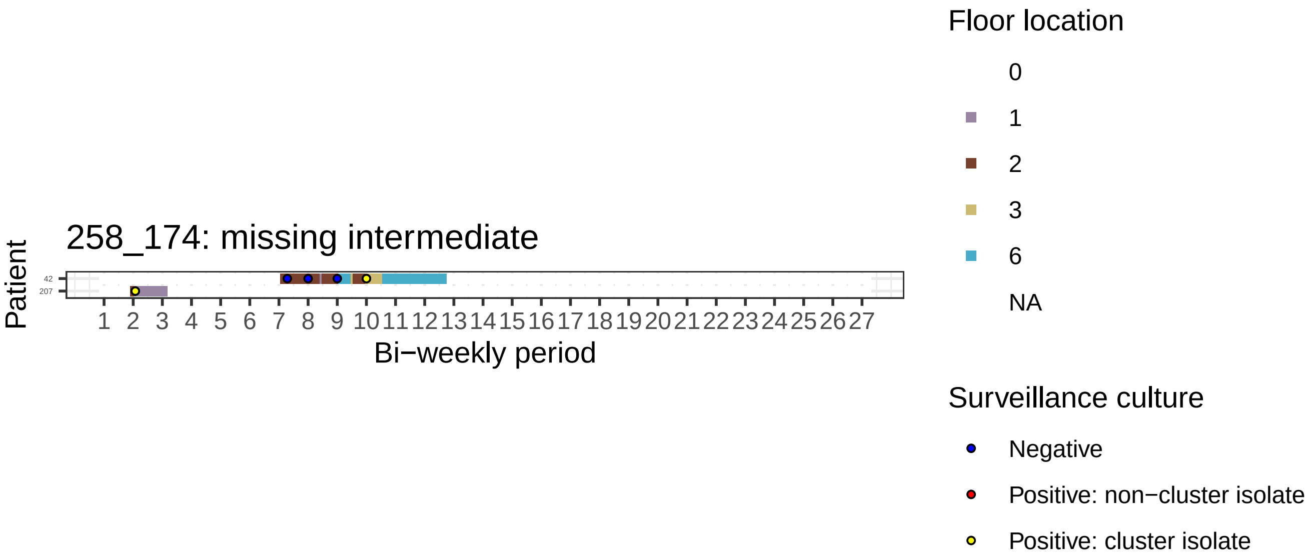
258_174: missing intermediate
Patient
42
207
1 2 3 4 5 6 7 8 9 10 11 12 13 14 15 16 17 18 19 20 21 22 23 24 25 26 27
Bi−weekly period
Floor location
0
1
2
3
6
NA
Surveillance culture
Negative
Positive: non−cluster isolate
Positive: cluster isolate

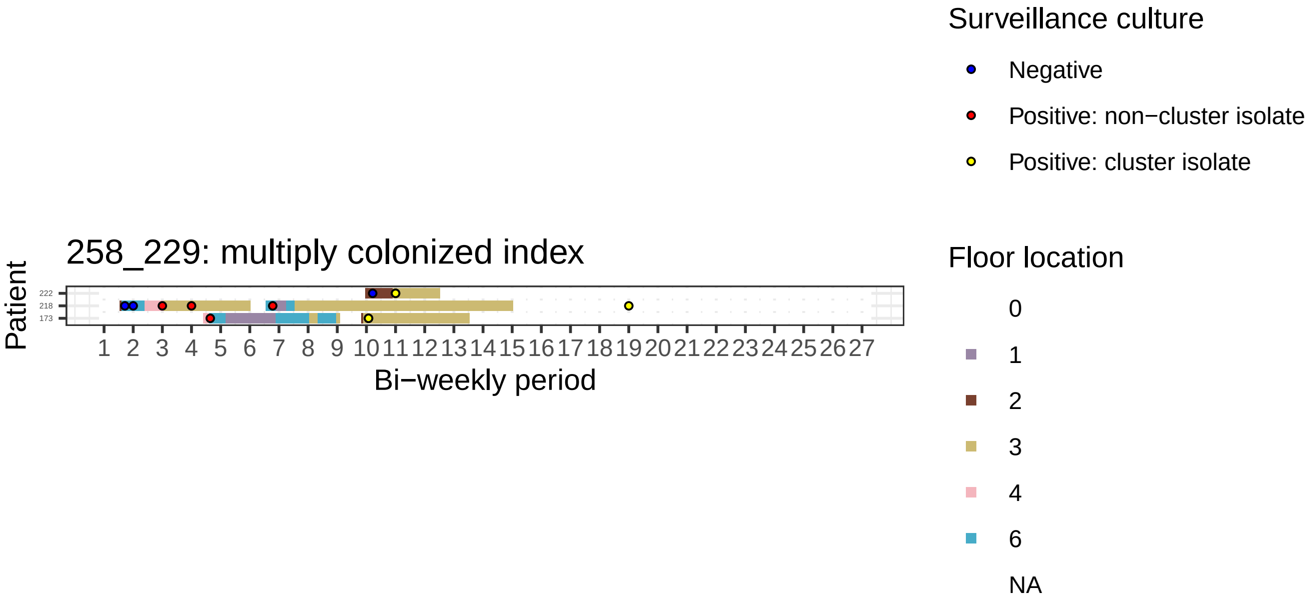
258_229: multiply colonized index
Patient
222
218
173
1 2 3 4 5 6 7 8 9 10 11 12 13 14 15 16 17 18 19 20 21 22 23 24 25 26 27
Bi−weekly period
Surveillance culture
Negative
Positive: non−cluster isolate
Positive: cluster isolate
Floor location
0
1
2
3
4
6
NA

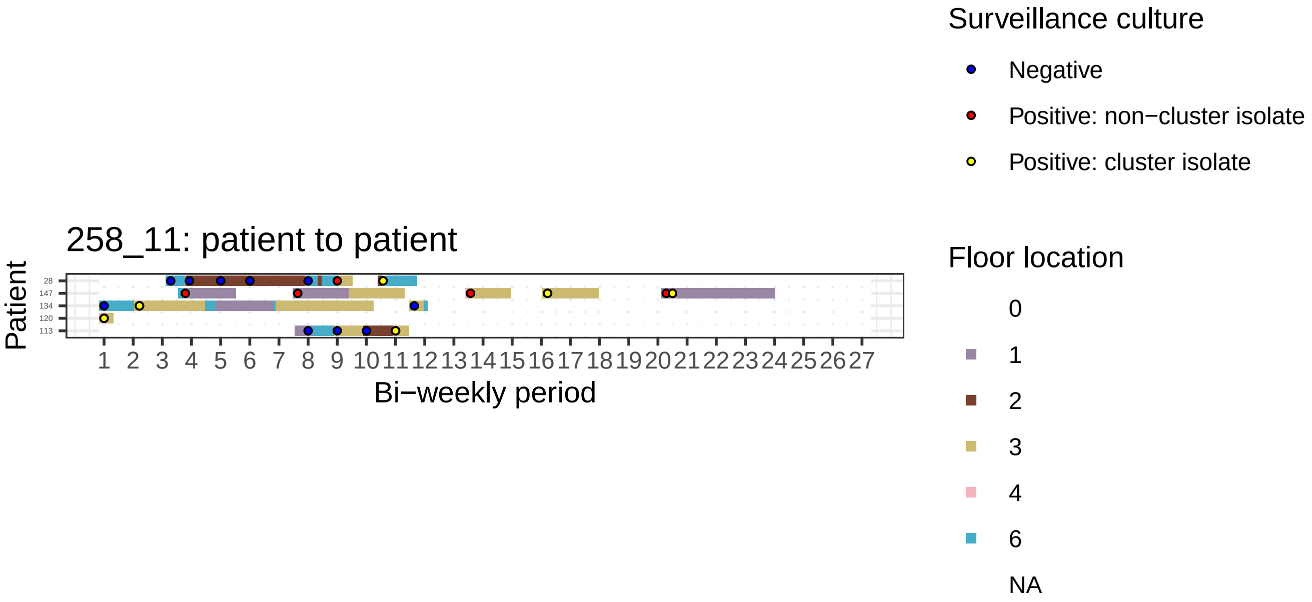

258_11: patient to patient
Patient
28
147
134
120
113
Bi−weekly period
Surveillance culture
Negative
Positive: non−cluster isolate
Positive: cluster isolate
Floor location
0
1
2
3
4
6
NA

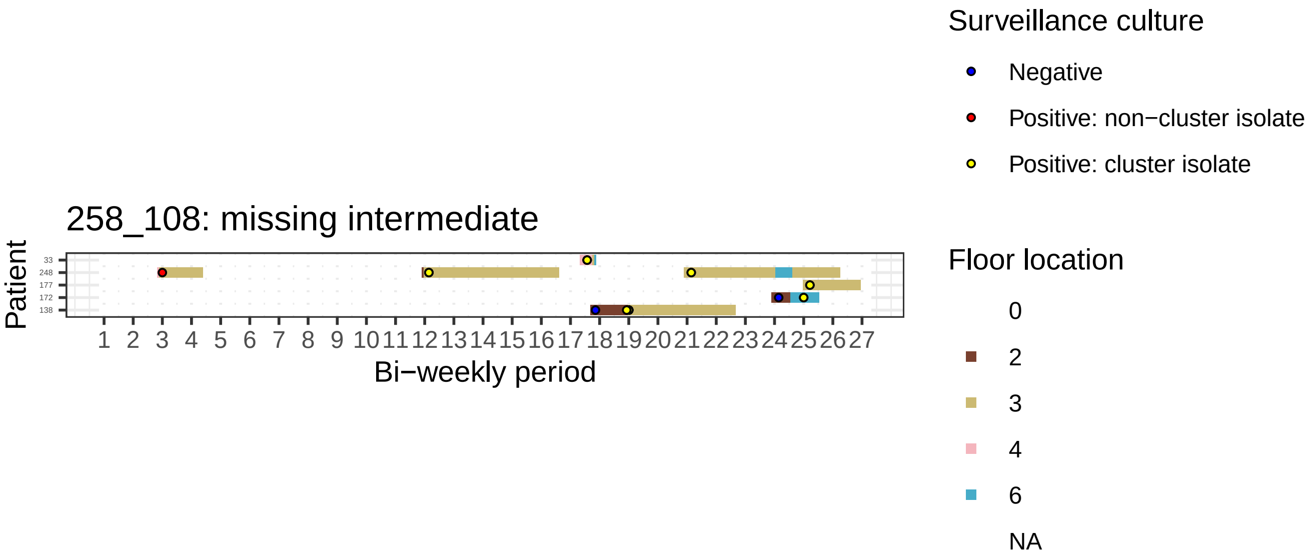
258_108: missing intermediate
Patient
33
248
177
172
138
1 2 3 4 5 6 7 8 9 10 11 12 13 14 15 16 17 18 19 20 21 22 23 24 25 26 27
Bi−weekly period
Surveillance culture
Negative
Positive: non−cluster isolate
Positive: cluster isolate
Floor location
0
2
3
4
6
NA

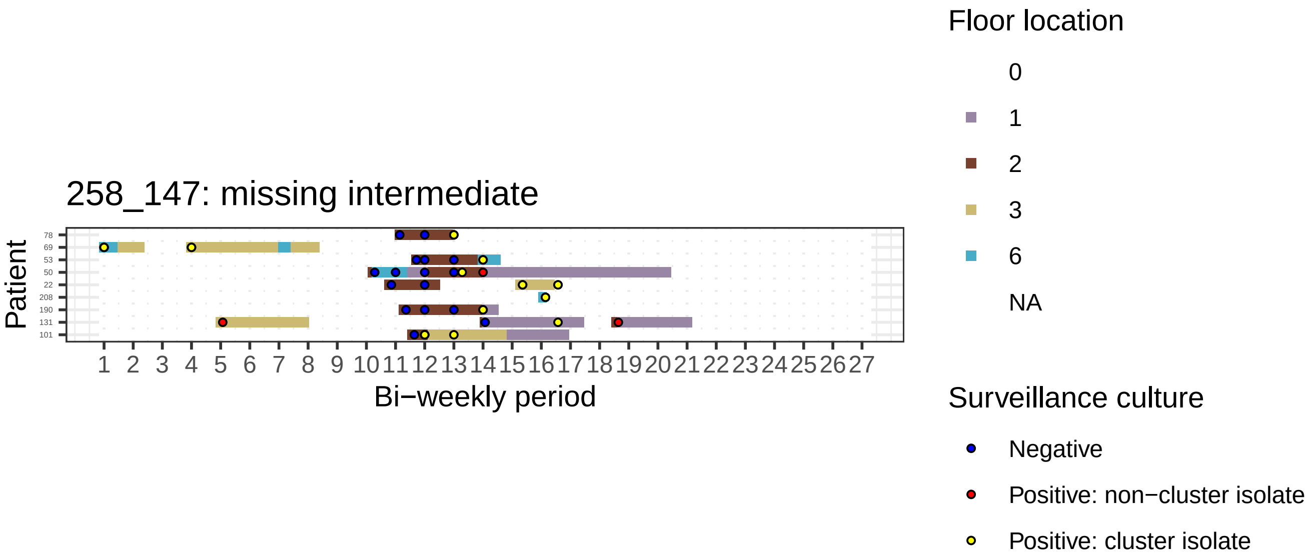

258_147: missing intermediate
Patient
78
69
53
50
22
208
190
131
101
1 2 3 4 5 6 7 8 9 10 11 12 13 14 15 16 17 18 19 20 21 22 23 24 25 26 27
Bi−weekly period
Floor location
0
1
2
3
6
NA
Surveillance culture
Negative
Positive: non−cluster isolate
Positive: cluster isolate

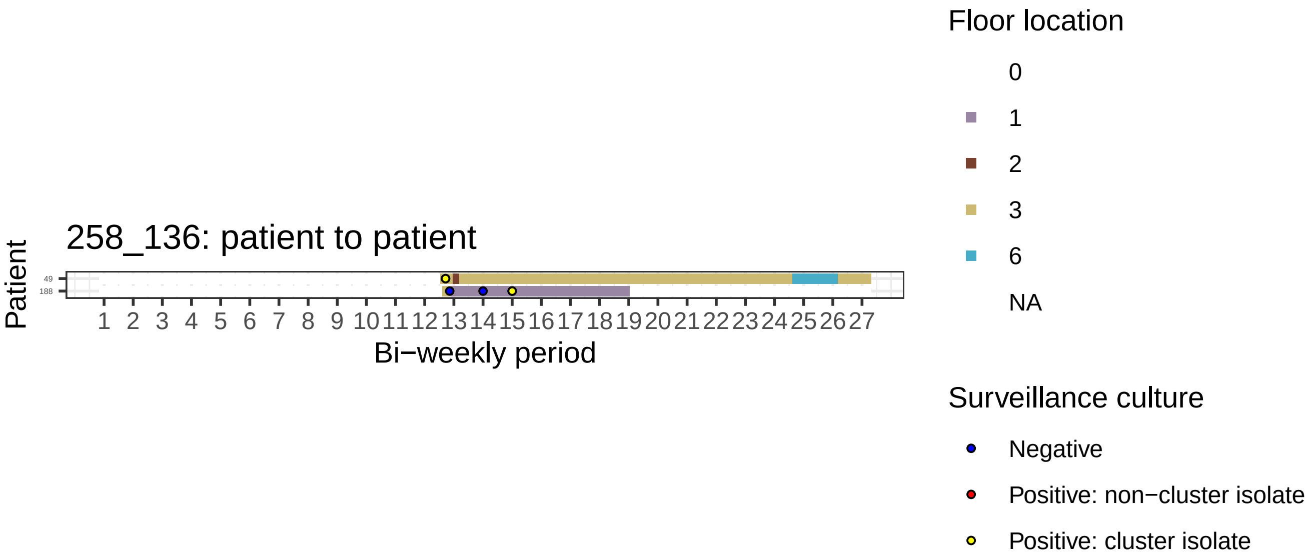

258_136: patient to patient
Patient
49
188
1 2 3 4 5 6 7 8 9 10 11 12 13 14 15 16 17 18 19 20 21 22 23 24 25 26 27
Bi−weekly period
Floor location
0
1
2
3
6
NA
Surveillance culture
Negative
Positive: non−cluster isolate
Positive: cluster isolate

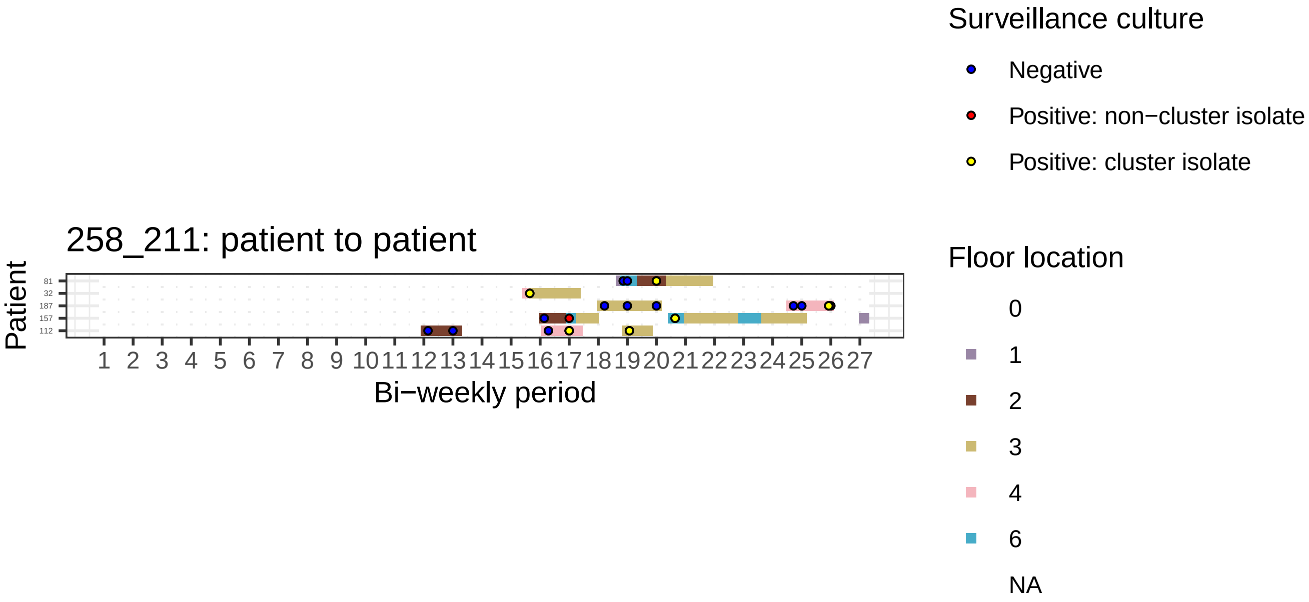

258_211: patient to patient
Patient
81
32
187
157
112
1 2 3 4 5 6 7 8 9 10 11 12 13 14 15 16 17 18 19 20 21 22 23 24 25 26 27
Bi−weekly period
Surveillance culture
Negative
Positive: non−cluster isolate
Positive: cluster isolate
Floor location
0
1
2
3
4
6
NA

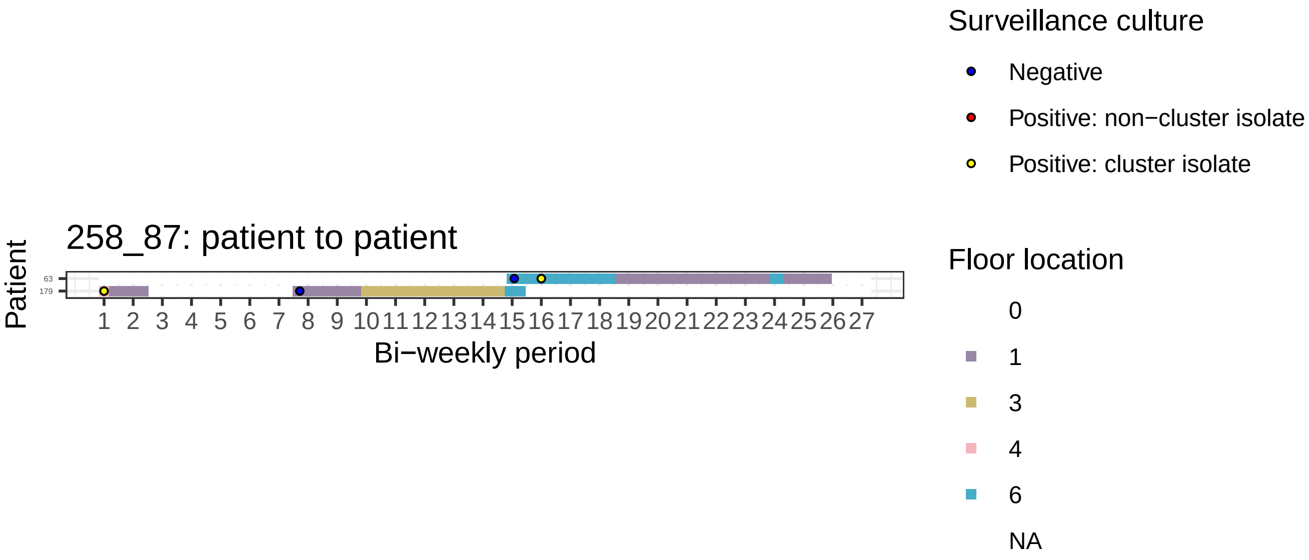
258_87: patient to patient
Patient
63
179
1 2 3 4 5 6 7 8 9 10 11 12 13 14 15 16 17 18 19 20 21 22 23 24 25 26 27
Bi−weekly period
Surveillance culture
Negative
Positive: non−cluster isolate
Positive: cluster isolate
Floor location
0
1
3
4
6
NA

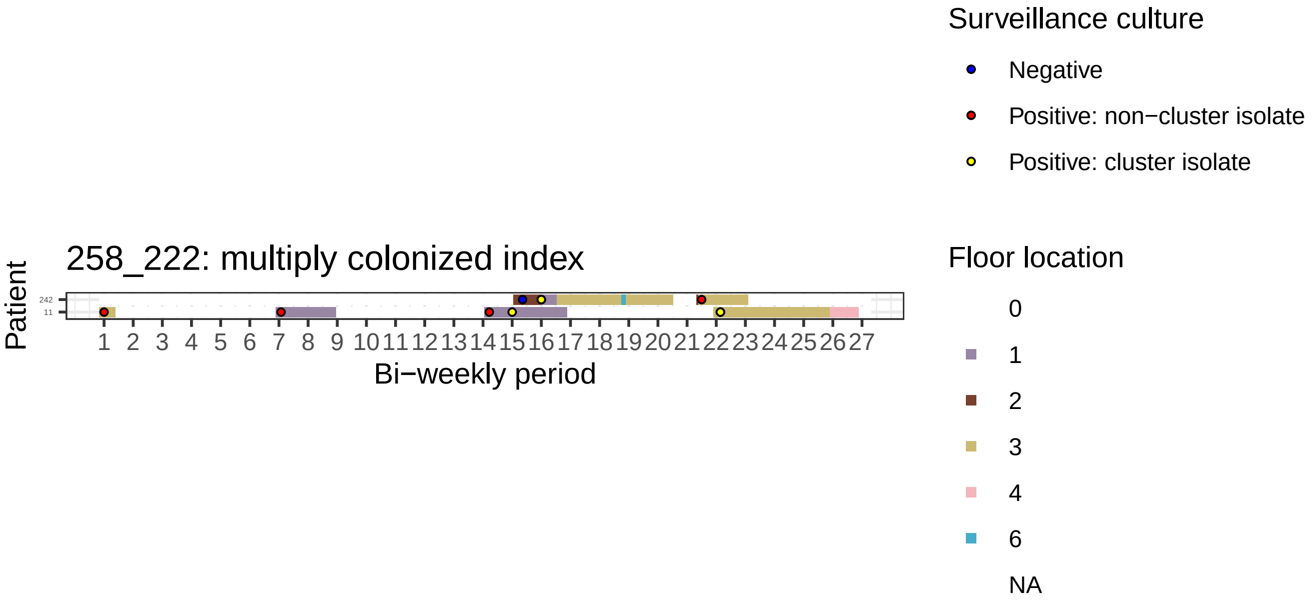

258_222: multiply colonized index
Patient
242
11
1 2 3 4 5 6 7 8 9 10 11 12 13 14 15 16 17 18 19 20 21 22 23 24 25 26 27
Bi−weekly period
Surveillance culture
Negative
Positive: non−cluster isolate
Positive: cluster isolate
Floor location
0
1
2
3
4
6
NA

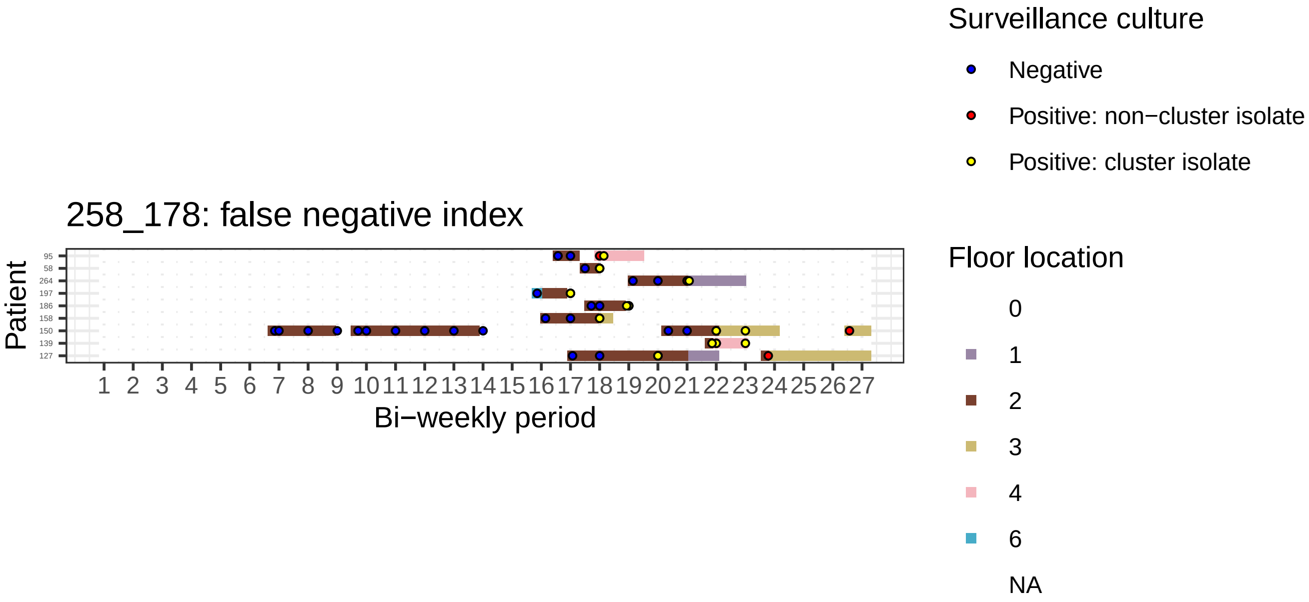
258_178: false negative index
Patient
95
58
264
197
186
158
150
139
127
1 2 3 4 5 6 7 8 9 10 11 12 13 14 15 16 17 18 19 20 21 22 23 24 25 26 27
Bi−weekly period
Surveillance culture
Negative
Positive: non−cluster isolate
Positive: cluster isolate
Floor location
0
1
2
3
4
6
NA

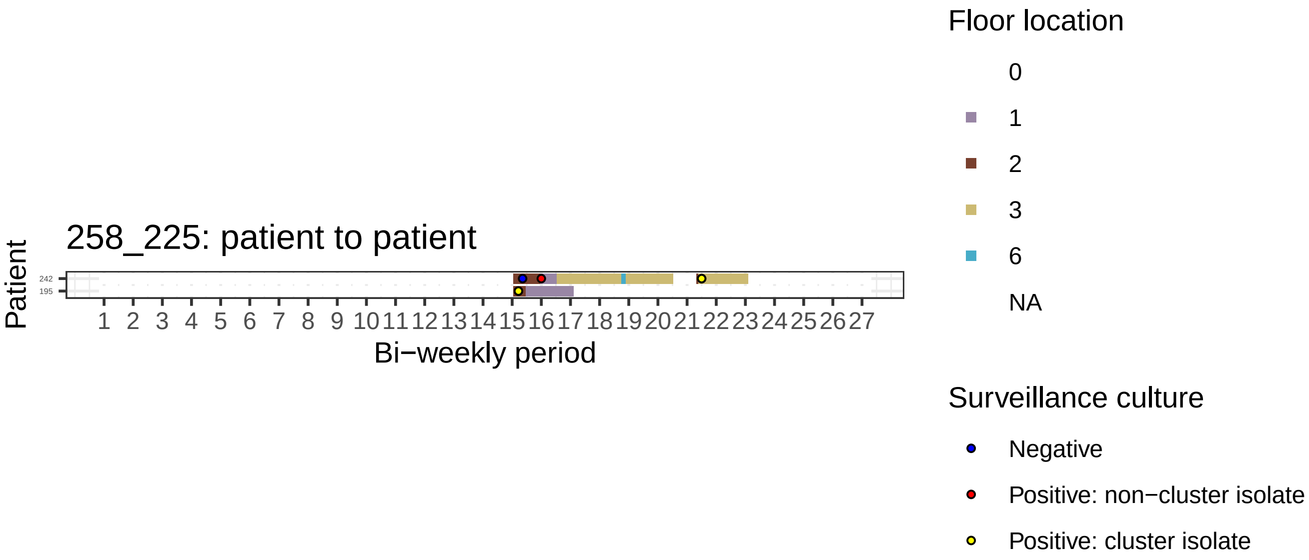

258_225: patient to patient
Patient
242
195
1 2 3 4 5 6 7 8 9 10 11 12 13 14 15 16 17 18 19 20 21 22 23 24 25 26 27
Bi−weekly period
Floor location
0
1
2
3
6
NA
Surveillance culture
Negative
Positive: non−cluster isolate
Positive: cluster isolate

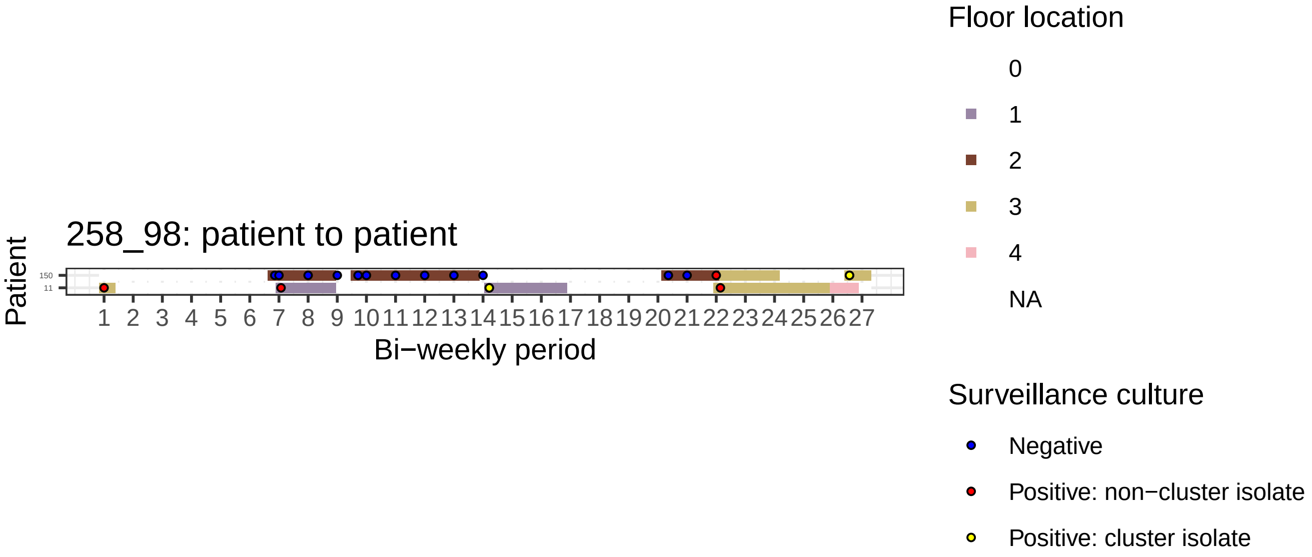

258_98: patient to patient
Patient
150
11
1 2 3 4 5 6 7 8 9 10 11 12 13 14 15 16 17 18 19 20 21 22 23 24 25 26 27
Bi−weekly period
Floor location
0
1
2
3
4
NA
Surveillance culture
Negative
Positive: non−cluster isolate
Positive: cluster isolate

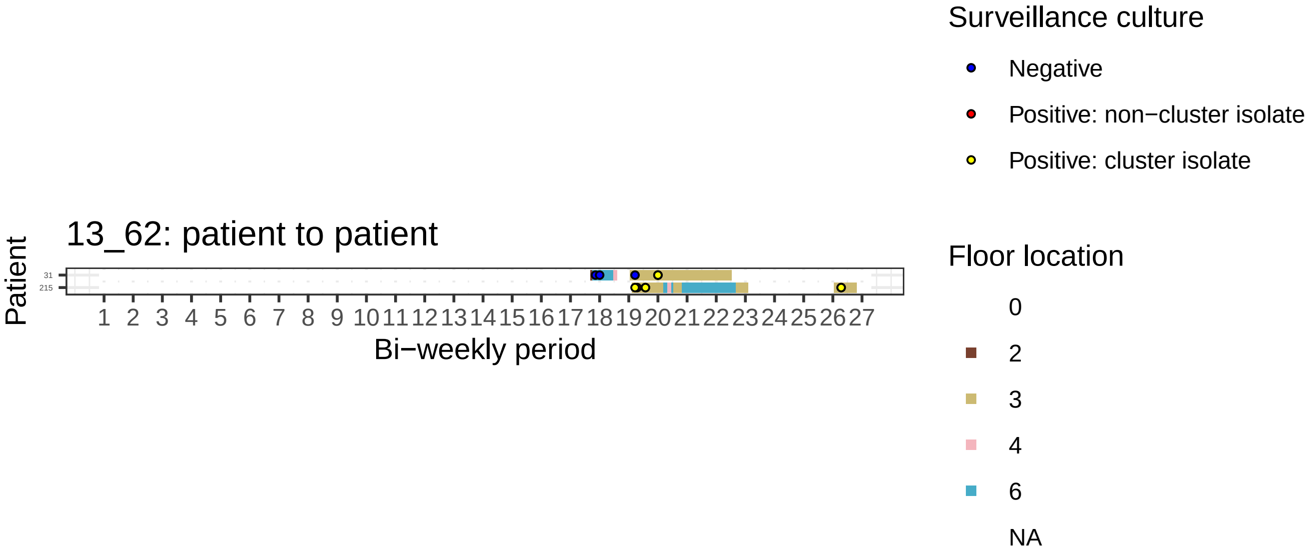
13_62: patient to patient
Patient
31
215
Bi−weekly period
Surveillance culture
Negative
Positive: non−cluster isolate
Positive: cluster isolate
Floor location
0
2
3
4
6
NA

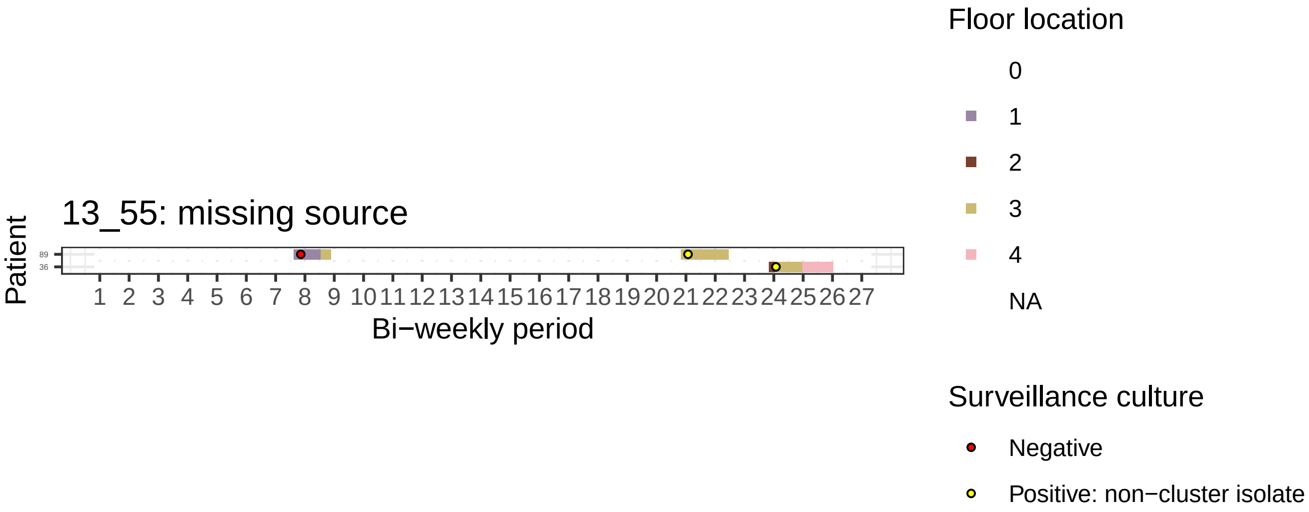

13_55: missing source
Patient
89
36
1 2 3 4 5 6 7 8 9 10 11 12 13 14 15 16 17 18 19 20 21 22 23 24 25 26 27
Bi−weekly period
Floor location
0
1
2
3
4
NA
Surveillance culture
Negative
Positive: non−cluster isolate

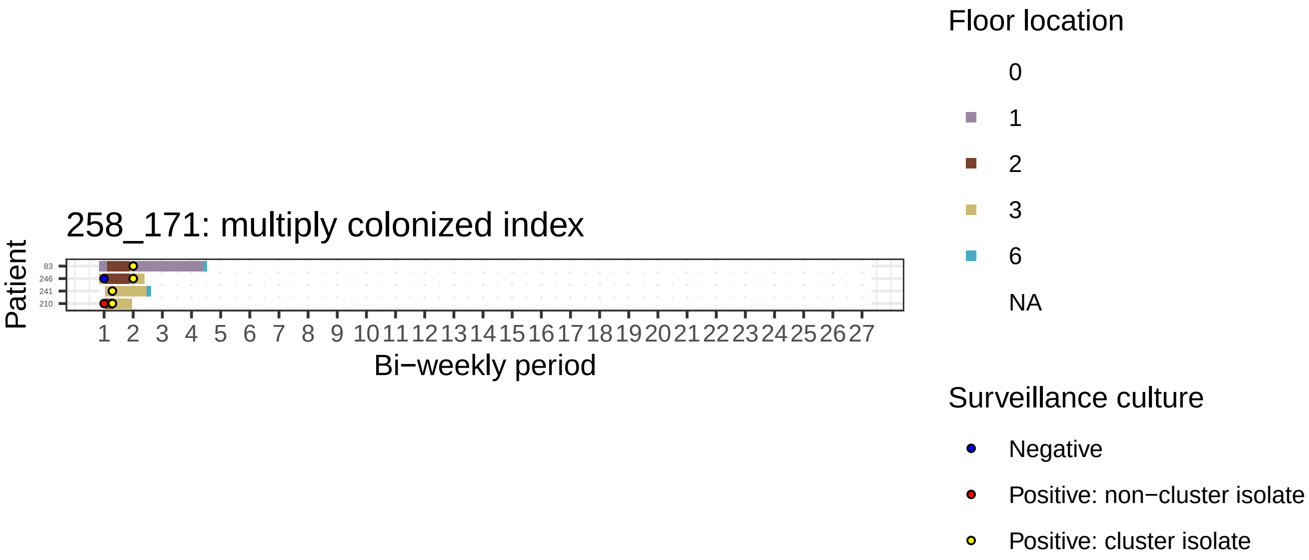
258_171: multiply colonized index
Patient
83
246
241
210
1 2 3 4 5 6 7 8 9 10 11 12 13 14 15 16 17 18 19 20 21 22 23 24 25 26 27
Bi−weekly period
Floor location
0
1
2
3
6
NA
Surveillance culture
Negative
Positive: non−cluster isolate
Positive: cluster isolate

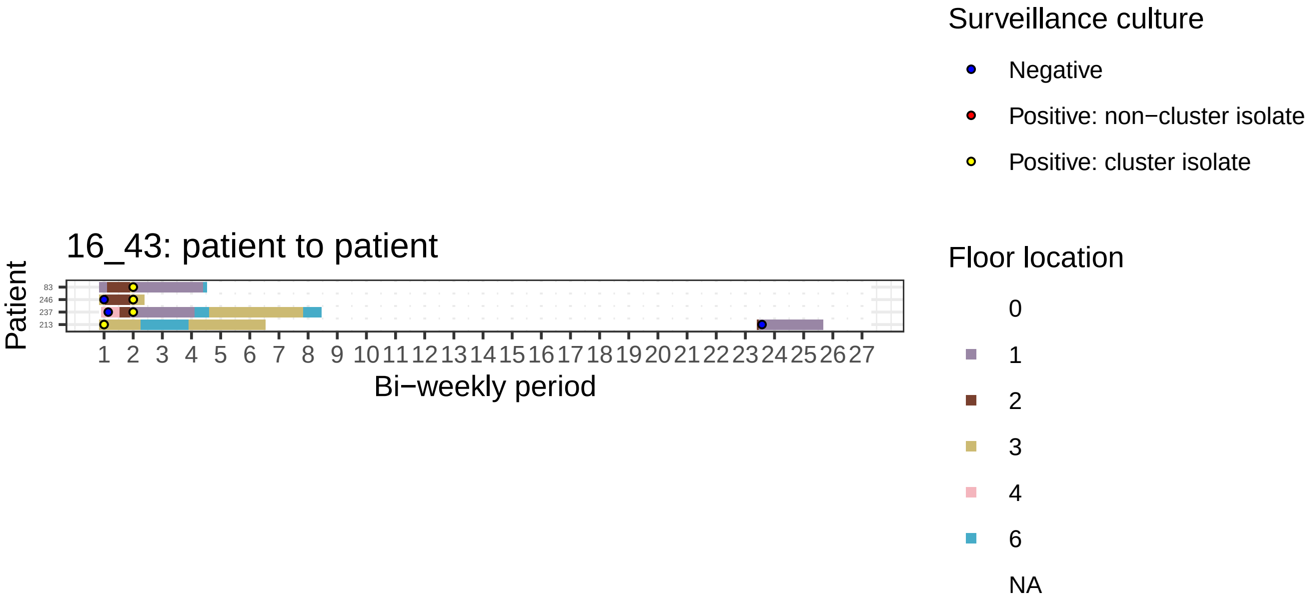

16_43: patient to patient
Patient
83
246
237
213
1 2 3 4 5 6 7 8 9 10 11 12 13 14 15 16 17 18 19 20 21 22 23 24 25 26 27
Bi−weekly period
Surveillance culture
Negative
Positive: non−cluster isolate
Positive: cluster isolate
Floor location
0
1
2
3
4
6
NA

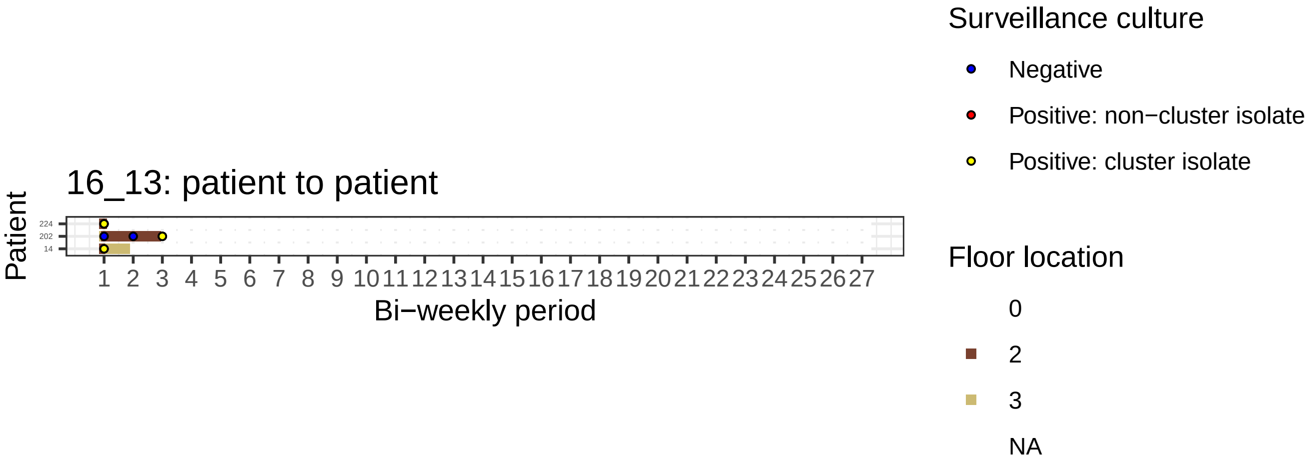
16_13: patient to patient
Patient
224
202
14
1 2 3 4 5 6 7 8 9 10 11 12 13 14 15 16 17 18 19 20 21 22 23 24 25 26 27
Bi−weekly period
Surveillance culture
Negative
Positive: non−cluster isolate
Positive: cluster isolate
Floor location
0
2
3
NA

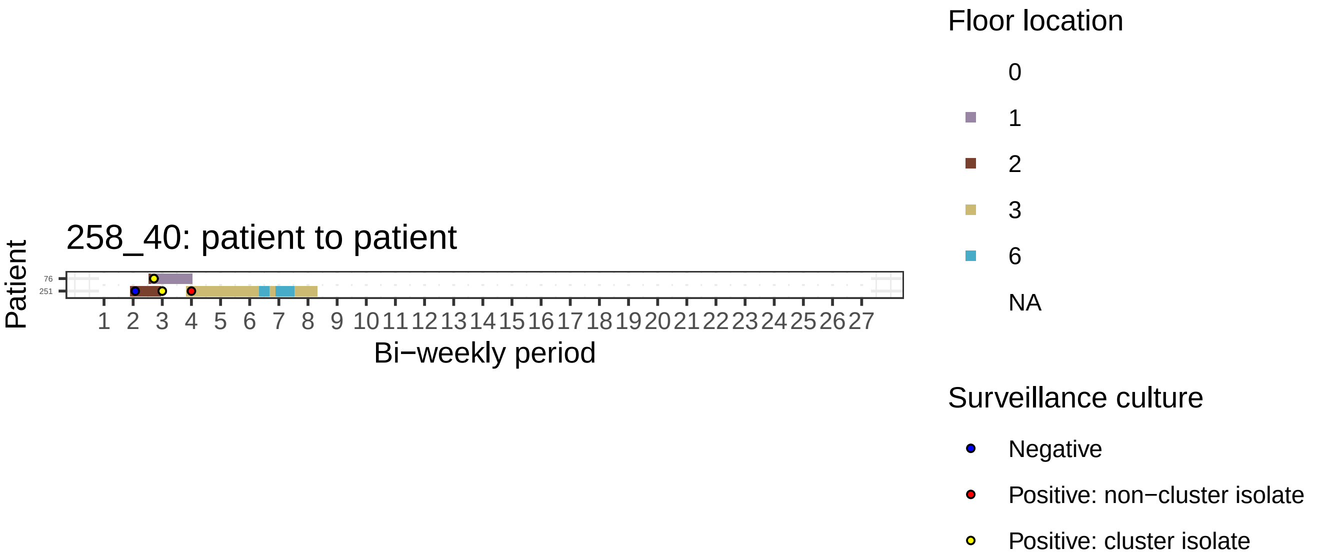

258_40: patient to patient
Patient
76
251
Bi−weekly period
Floor location
0
1
2
3
6
NA
Surveillance culture
Negative
Positive: non−cluster isolate
Positive: cluster isolate

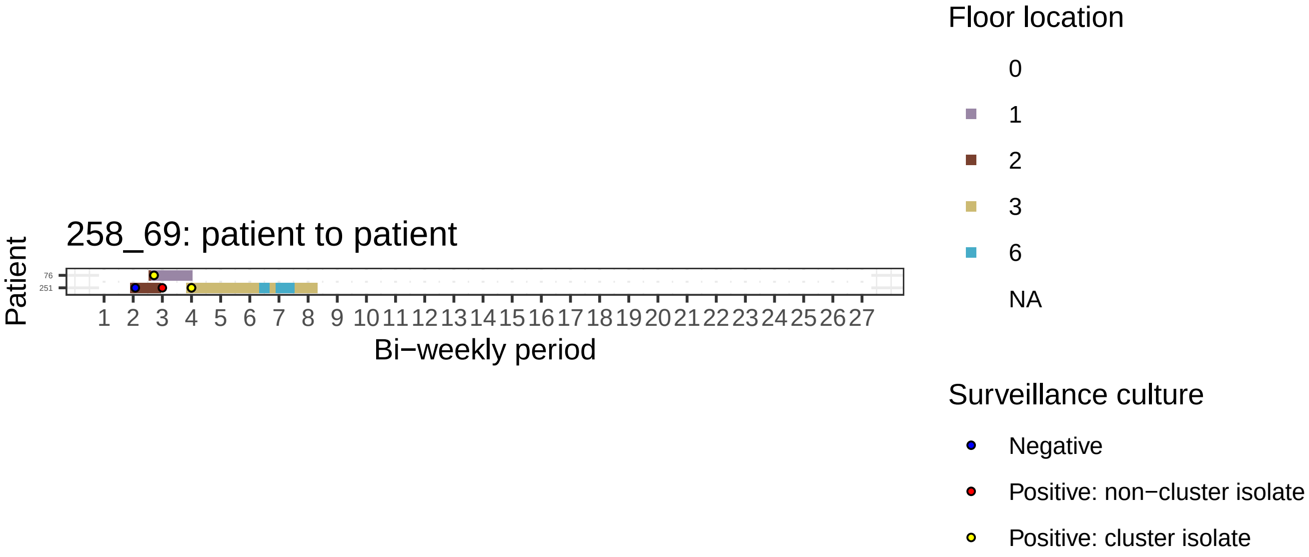

258_69: patient to patient
Patient
76
251
Bi−weekly period
Floor location
0
1
2
3
6
NA
Surveillance culture
Negative
Positive: non−cluster isolate
Positive: cluster isolate

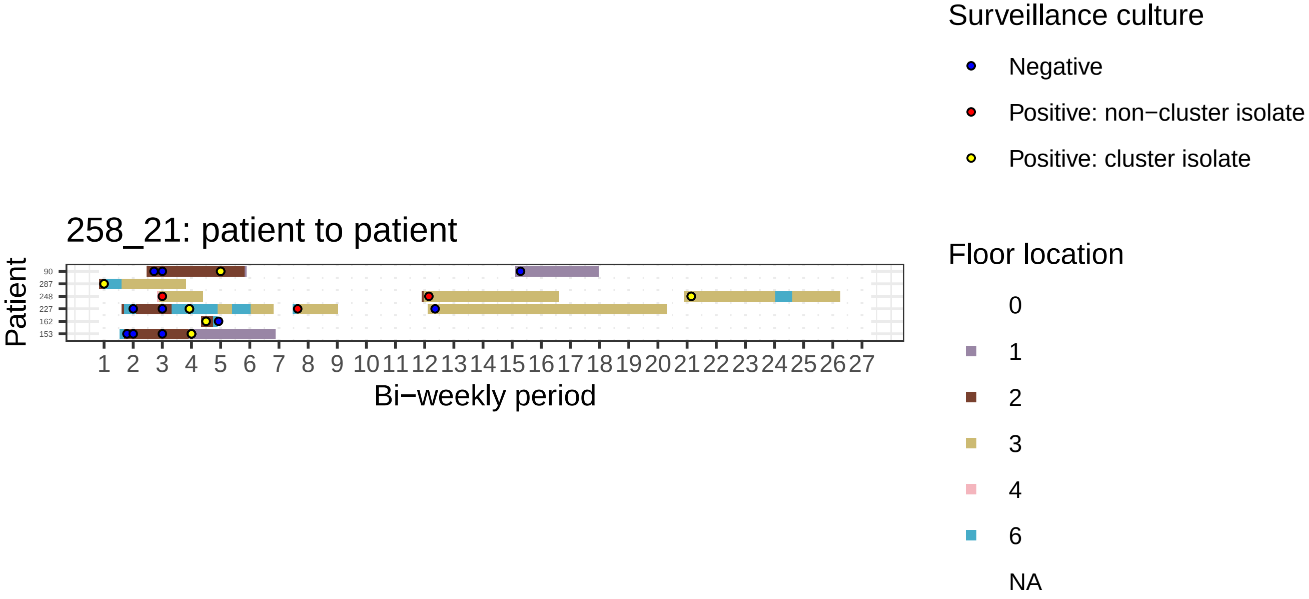

258_21: patient to patient
Patient
90
287
248
227
162
153
1 2 3 4 5 6 7 8 9 10 11 12 13 14 15 16 17 18 19 20 21 22 23 24 25 26 27
Bi−weekly period
Surveillance culture
Negative
Positive: non−cluster isolate
Positive: cluster isolate
Floor location
0
1
2
3
4
6
NA

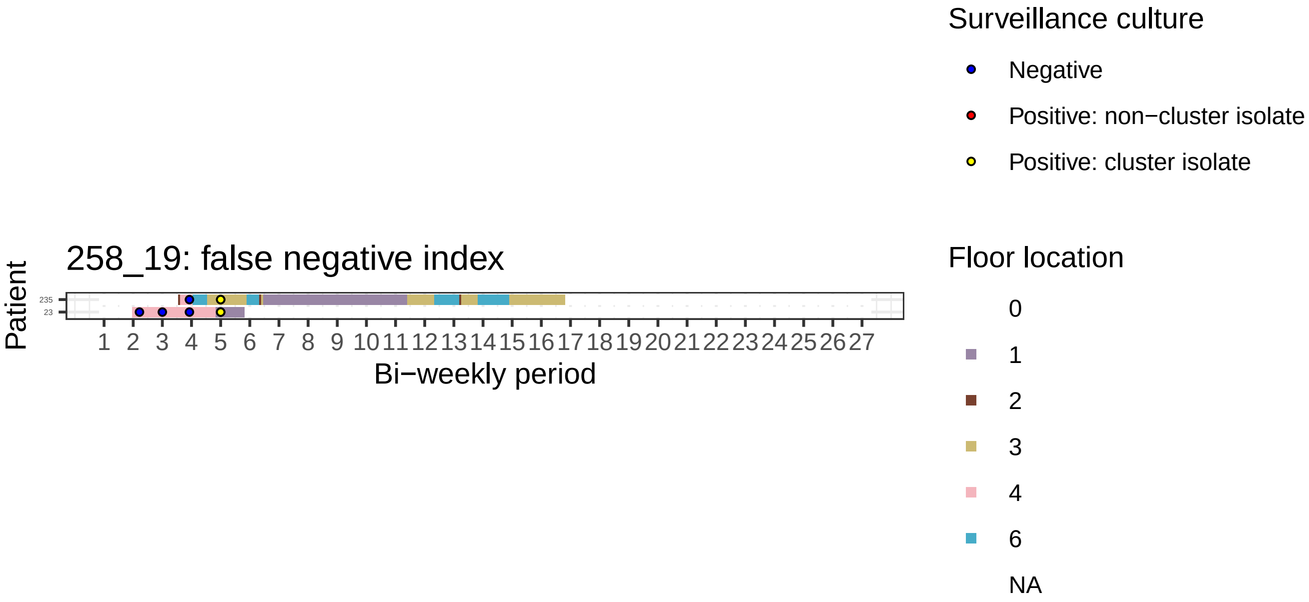

258_19: false negative index
Patient
235
23
1 2 3 4 5 6 7 8 9 10 11 12 13 14 15 16 17 18 19 20 21 22 23 24 25 26 27
Bi−weekly period
Surveillance culture
Negative
Positive: non−cluster isolate
Positive: cluster isolate
Floor location
0
1
2
3
4
6
NA

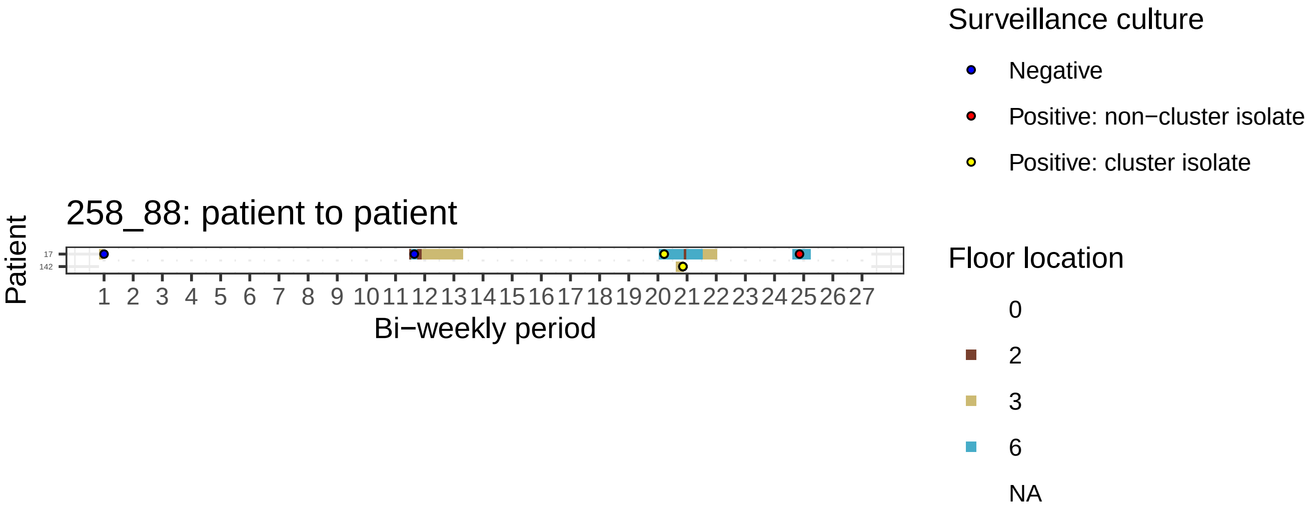
258_88: patient to patient
Patient
17
142
1 2 3 4 5 6 7 8 9 10 11 12 13 14 15 16 17 18 19 20 21 22 23 24 25 26 27
Bi−weekly period
Surveillance culture
Negative
Positive: non−cluster isolate
Positive: cluster isolate
Floor location
0
2
3
6
NA

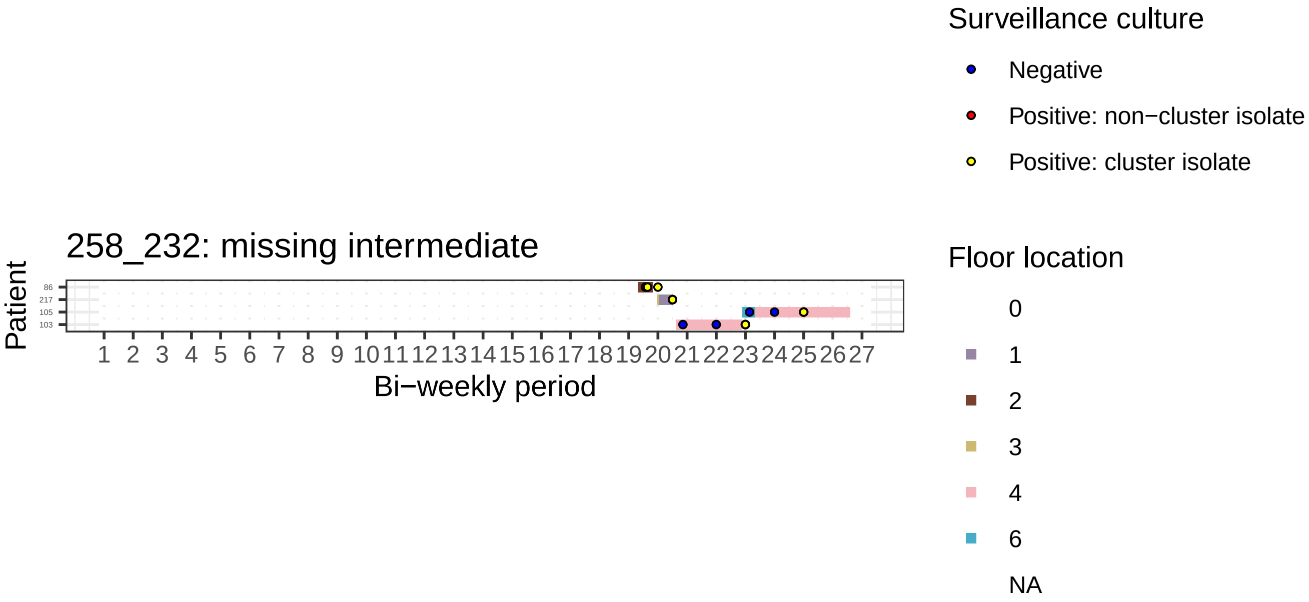
258_232: missing intermediate
Patient
86
217
105
103
1 2 3 4 5 6 7 8 9 10 11 12 13 14 15 16 17 18 19 20 21 22 23 24 25 26 27
Bi−weekly period
Surveillance culture
Negative
Positive: non−cluster isolate
Positive: cluster isolate
Floor location
0
1
2
3
4
6
NA

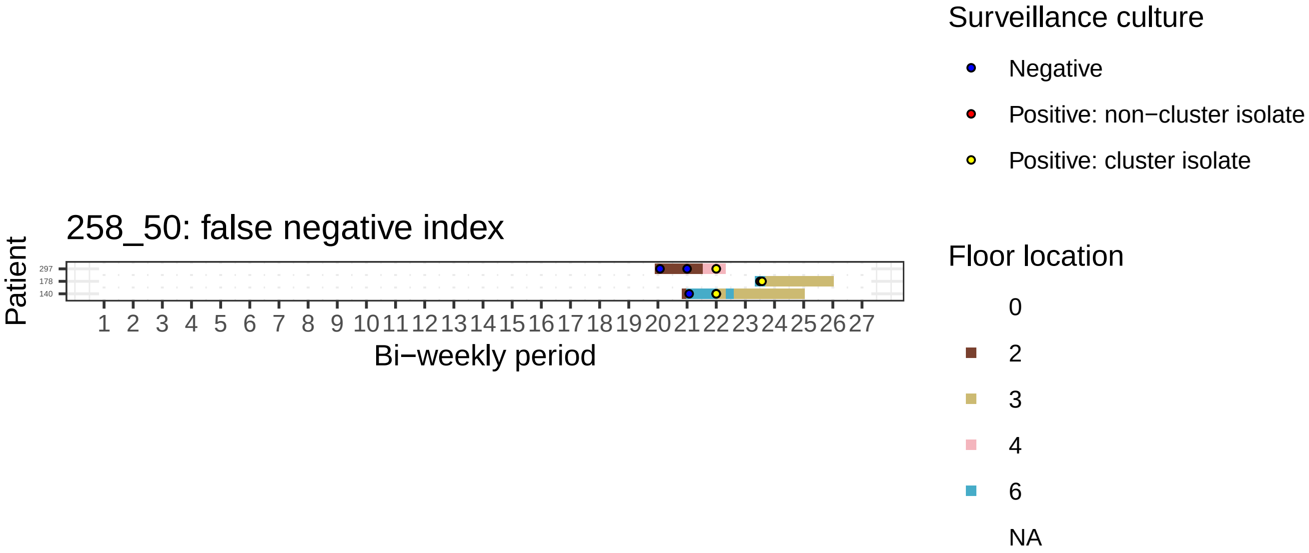
258_50: false negative index
Patient
297
178
140
1 2 3 4 5 6 7 8 9 10 11 12 13 14 15 16 17 18 19 20 21 22 23 24 25 26 27
Bi−weekly period
Surveillance culture
Negative
Positive: non−cluster isolate
Positive: cluster isolate
Floor location
0
2
3
4
6
NA

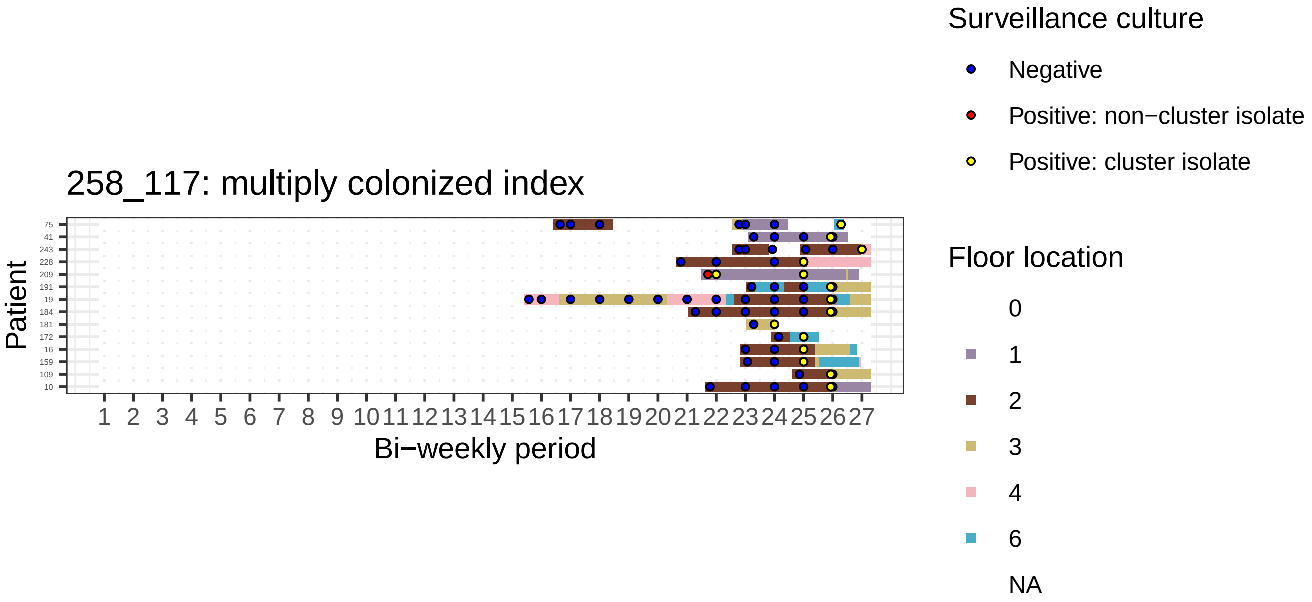

258_117: multiply colonized index
Patient
75
41
243
228
209
191
19
184
181
172
16
159
109
10
1 2 3 4 5 6 7 8 9 10 11 12 13 14 15 16 17 18 19 20 21 22 23 24 25 26 27
Bi−weekly period
Surveillance culture
Negative
Positive: non−cluster isolate
Positive: cluster isolate
Floor location
0
1
2
3
4
6
NA

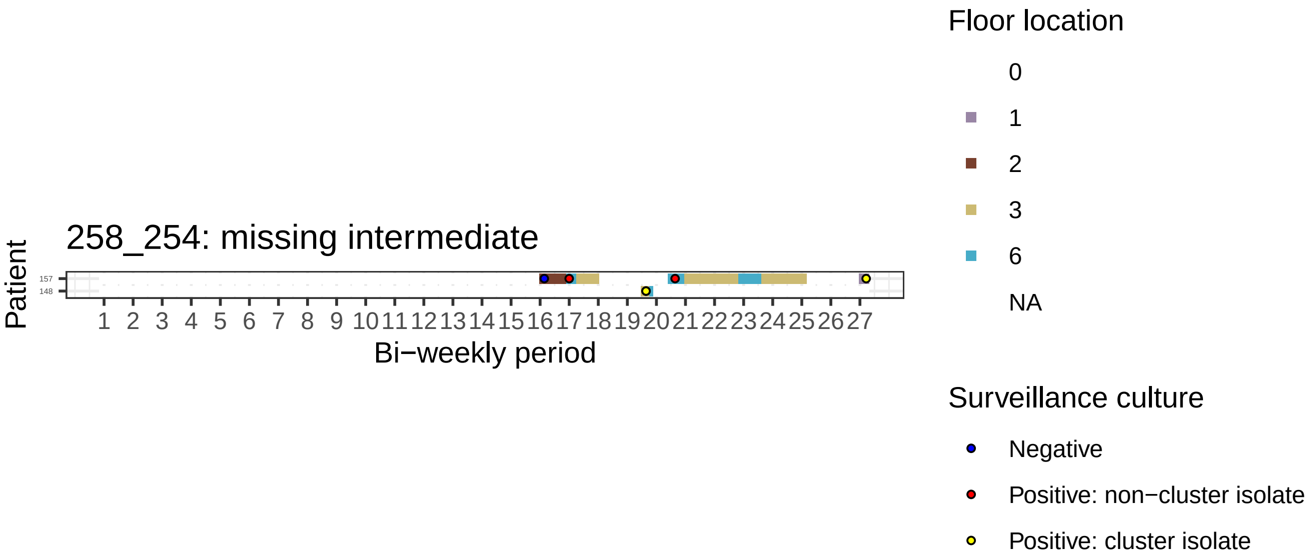
258_254: missing intermediate
Patient
157
148
1 2 3 4 5 6 7 8 9 10 11 12 13 14 15 16 17 18 19 20 21 22 23 24 25 26 27
Bi−weekly period
Floor location
0
1
2
3
6
NA
Surveillance culture
Negative
Positive: non−cluster isolate
Positive: cluster isolate

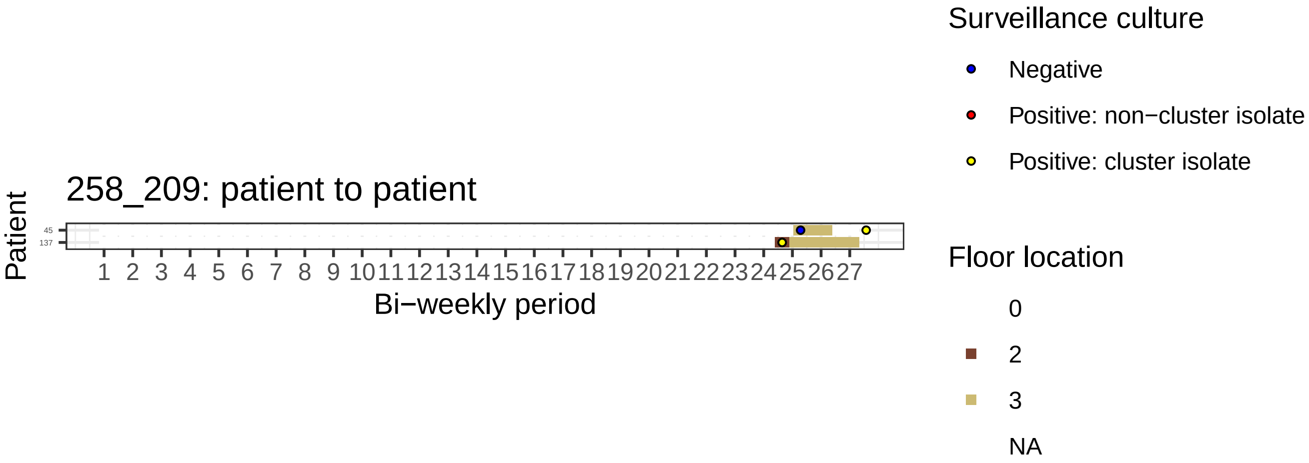

258_209: patient to patient
Patient
45
137
1 2 3 4 5 6 7 8 9 10 11 12 13 14 15 16 17 18 19 20 21 22 23 24 25 26 27
Bi−weekly period
Surveillance culture
Negative
Positive: non−cluster isolate
Positive: cluster isolate
Floor location
0
2
3
NA

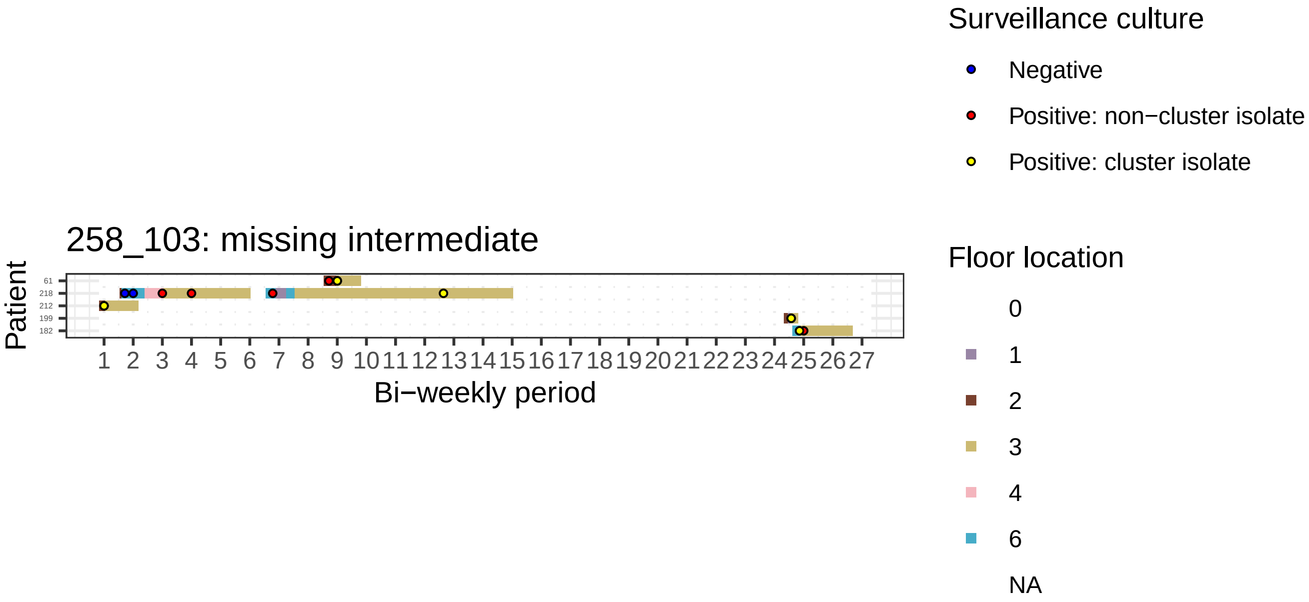
258_103: missing intermediate
Patient
61
218
212
199
182
1 2 3 4 5 6 7 8 9 10 11 12 13 14 15 16 17 18 19 20 21 22 23 24 25 26 27
Bi−weekly period
Surveillance culture
Negative
Positive: non−cluster isolate
Positive: cluster isolate
Floor location
0
1
2
3
4
6
NA

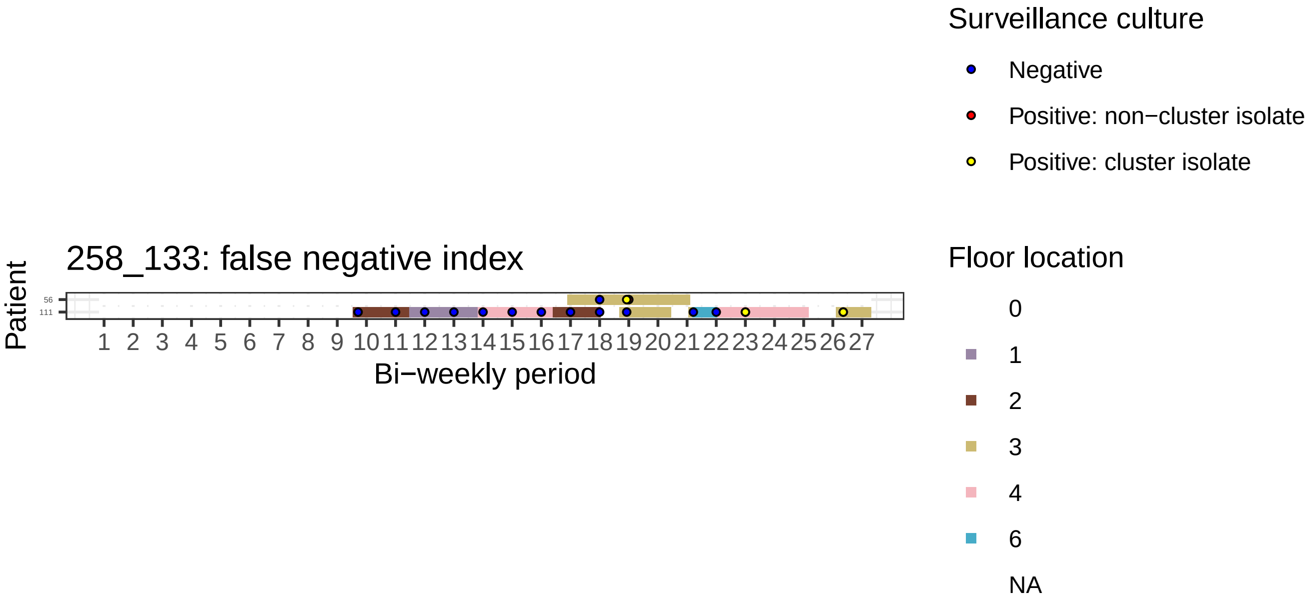
258_133: false negative index
Patient
56
111
1 2 3 4 5 6 7 8 9 10 11 12 13 14 15 16 17 18 19 20 21 22 23 24 25 26 27
Bi−weekly period
Surveillance culture
Negative
Positive: non−cluster isolate
Positive: cluster isolate
Floor location
0
1
2
3
4
6
NA

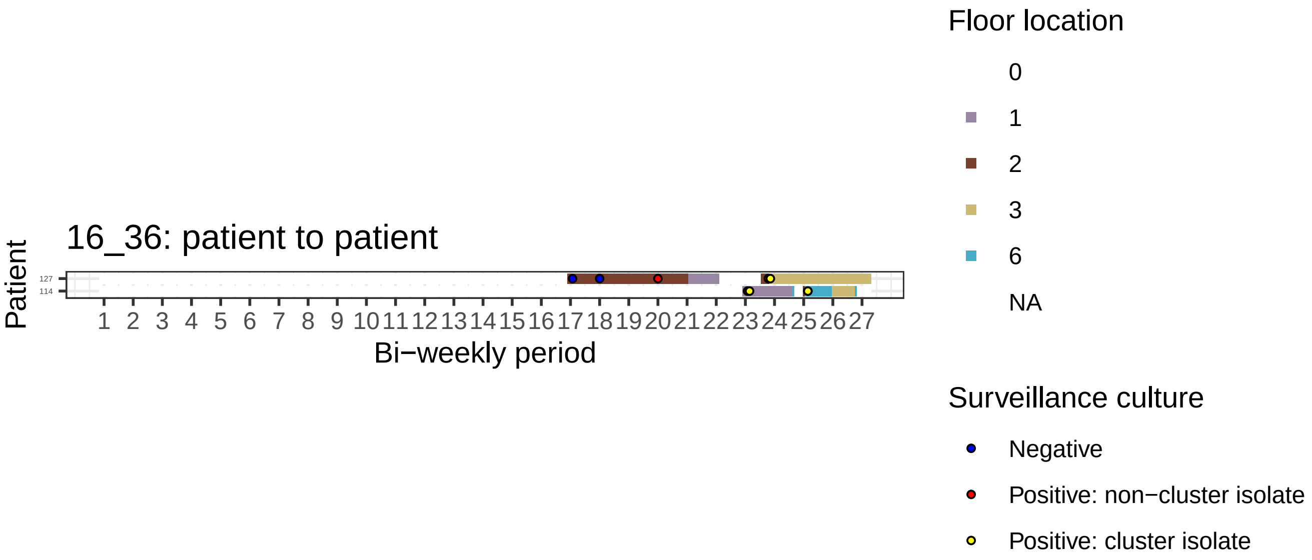
16_36: patient to patient
Patient
127
114
1 2 3 4 5 6 7 8 9 10 11 12 13 14 15 16 17 18 19 20 21 22 23 24 25 26 27
Bi−weekly period
Floor location
0
1
2
3
6
NA
Surveillance culture
Negative
Positive: non−cluster isolate
Positive: cluster isolate

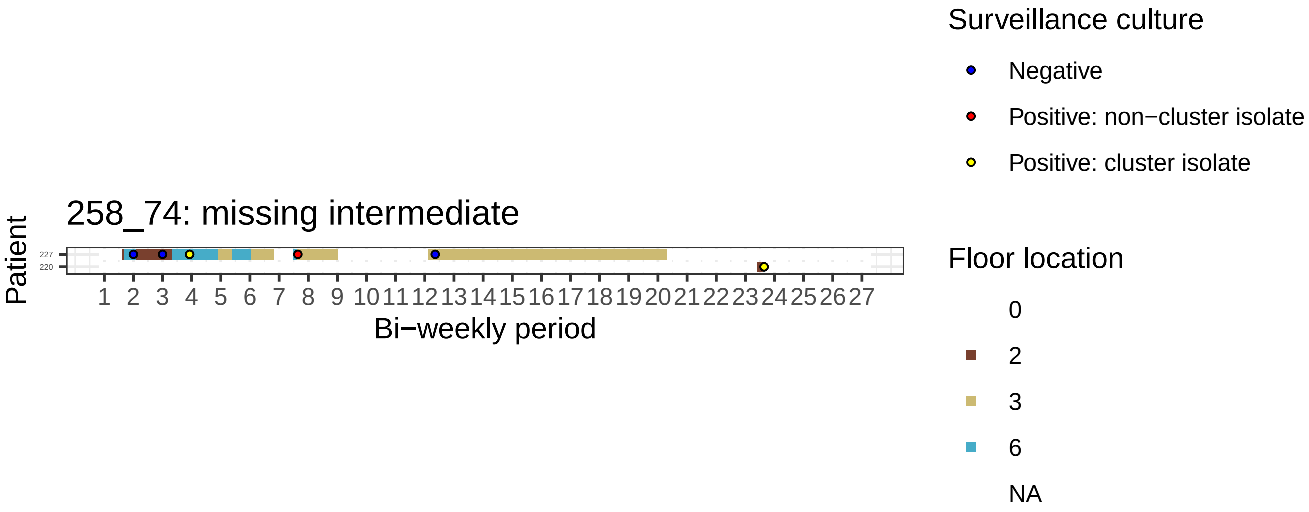
258_74: missing intermediate
Patient
227
220
Bi−weekly period
Surveillance culture
Negative
Positive: non−cluster isolate
Positive: cluster isolate
Floor location
0
2
3
6
NA

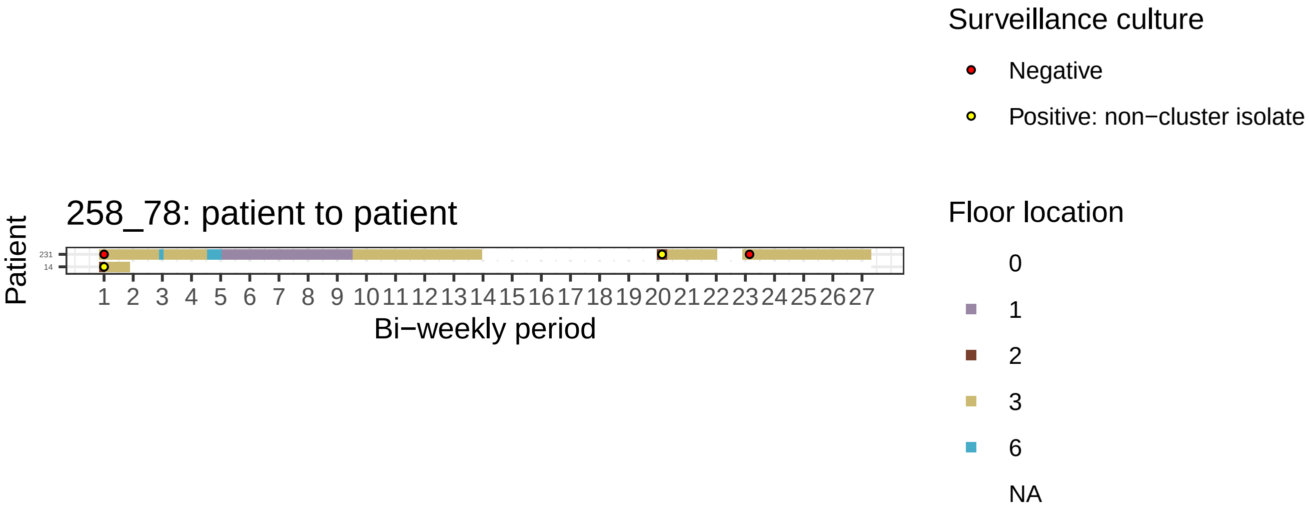

258_78: patient to patient
Patient
231
14
1 2 3 4 5 6 7 8 9 10 11 12 13 14 15 16 17 18 19 20 21 22 23 24 25 26 27
Bi−weekly period
Surveillance culture
Negative
Positive: non−cluster isolate
Floor location
0
1
2
3
6
NA

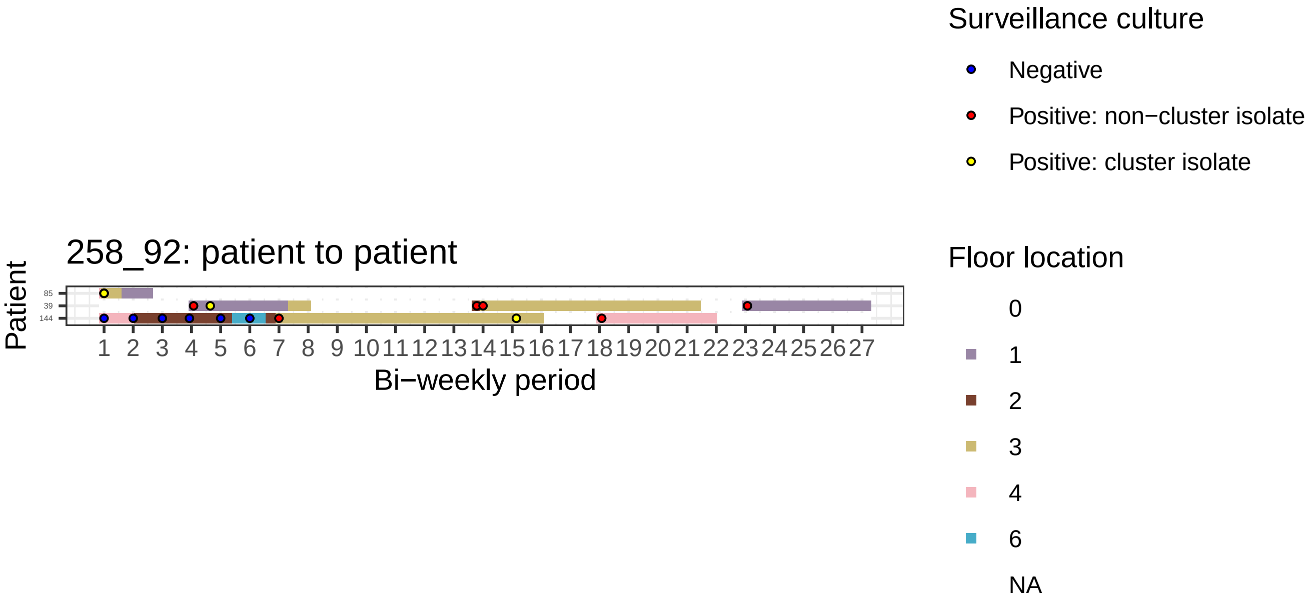

258_92: patient to patient
Patient
85
39
144
1 2 3 4 5 6 7 8 9 10 11 12 13 14 15 16 17 18 19 20 21 22 23 24 25 26 27
Bi−weekly period
Surveillance culture
Negative
Positive: non−cluster isolate
Positive: cluster isolate
Floor location
0
1
2
3
4
6
NA
